# RISK FACTORS FOR EARLY NATURAL MENOPAUSE: EVIDENCE FROM THE 1958 AND 1970 BRITISH BIRTH COHORTS

**DOI:** 10.1101/2021.09.12.21263444

**Authors:** Darina Peycheva, Alice Sullivan, Rebecca Hardy, Alex Bryson, Gabriella Conti, George Ploubidis

**Author notes:** Corresponding Author, DP is a corresponding author. All authors conceived and designed the study. DP performed the statistical analysis. All authors contributed to interpretation of data, revised the manuscript and approved the final version before submission.

## Abstract

Using data from two generations of British women followed from birth through childhood and into adulthood, we investigate risk factors for the onset of natural menopause before the age of 45 (known as early menopause). We focus on key stages during the life course to understand when risk factors are particularly harmful. We find that earlier cessation of menstruation is influenced by circumstances at birth. Women born in lower social class families, whose mother smoked during the pregnancy or who were short-term breastfed (one month or less) were more likely to undergo menopause before 45. Early menopause is also associated with poorer cognitive ability and smoking in childhood. Adult health behaviour also matters. Smoking is positively correlated with early menopause, while regular exercise (one to several times a week) and moderate frequency of alcohol drinking (one to three times a month) in women’s early thirties are associated with a reduced risk of early menopause. The occurrence of gynaecological problems by women’s early thirties is also linked to early menopause. We note that some of these factors (e.g. health behaviours) are modifiable and thus the risks may be preventable.

## 1. Introduction

Menopause signifies the permanent cessation of the ovarian function and the end of a woman’s reproductive period. Natural menopause is recognized to have occurred after 12 consecutive months of amenorrhea for which there is no other obvious pathological or physiological cause. It occurs with the final menstrual period (FMP), which is known with certainty only in retrospect a year or more after the event (WHO 1996). It usually occurs between 45 and 55 years of age with an average age at menopause estimated between 50 and 52 years though there are slight variations between ethnic groups (Mishra 2019). However, between 1 and 3% experience menopause before the age of 40, and between 5 and 10% experience menopause between the ages of 40 and 44 (Mishra 2019; Shifren et al 2014). Here, we will follow the convention of referring to the onset of menopause before age 45 as *early menopause*^*1*^.

Women with early menopause experience an extended period with loss of ovarian function and oestrogen deficiency and have an increased risk of cardiovascular disease (Zhu et al 2020a, 2020b), osteoporosis (Gallagher 2007), type 2 diabetes (Anagnostis et al 2019), premature decline in cognitive function (Kok et al 2006), decreased life expectancy and increased all-cause mortality (Mishra 2019; Gold 2011).

Past research suggests that the onset of menopause is strongly influenced by genetics (de Bruin et al 2001; Treloar et al 1998), but non-genetic factors can also play a role (Kok et al 2005; van Asselt et al 2004). However, there is little consensus on the influence of non-genetic factors on the timing of the menopause. Cigarette smoking has consistently been associated with early menopause, but the influence of several other factors, such as reproductive health, health behaviour (e.g. alcohol consumption, exercise), and socioeconomic circumstances, remains unclear. Consequently, it is critically important to establish the factors associated with early menopause, especially those that are modifiable so that appropriate preventive health strategies can be considered.

In addition, a life course approach (Mishra et al 2013; Hardy and Kuh 2005) has indicated that there are early life factors which may be associated with early menopause. The life course approach enables identification of critical or sensitive periods during life where exposure to a particular risk factor may have most effect and may thus provide information on timely interventions (Jacob et al 2017). A life course approach is relevant for investigating age at menopause because a woman’s ovarian reserve declines gradually from about several million follicles post-conception to about one million at birth, to 500,000 when menarche begins, decreasing to around a thousand prior to menopause (Harlow 2000). This indicates that factors operating both before and after birth may play a role in the follicle pool depletion and identifying such factors may help in understanding of biological process of menopause. Previous studies have noted that fetal and early life factors such as fetal growth and early life nutrition, early life stress and cognition, and childhood socioeconomic circumstances may play a role in the timing of menopause, but few studies have prospectively collected measures of multiple factors from early life alongside later life exposures (Hardy and Kuh 2005; Hardy and Kuh 2002).

It has been suggested that the lack of consensus on factors affecting the timing of menopause could be explained by methodological differences, e.g. differences in study populations and designs, differences in definition of menopause, and varying use of controls in the analyses (Gold 2011, WHO 1996). Analysis of large-scale prospective studies (Gold 2011) can help resolve this lack of consensus. This study investigates risk factors from across the life course for early menopause using data from two cohort studies of British women born in 1958 and 1970 and followed from birth through to childhood and adulthood (See: http://www.cls.ucl.ac.uk/ncds; http://www.cls.ucl.ac.uk/bcs70). Using data from two studies increases the numbers experiencing menopause before the age of 45 which provides greater statistical power, while also allowing assessment of consistency of associations across both generations. We investigate whether early life exposures (e.g. maternal smoking in pregnancy, maternal age at birth, birthweight, gestational age, and breastfeeding), reproductive characteristics (e.g. age at menarche, gynaecological problems, contraceptive use, and live births), health behaviour (smoking, alcohol consumption, and exercise), socioeconomic characteristics (e.g. father’s social class at birth and adult social class), cognitive ability, psychological distress and BMI are associated with early menopause. We focus on exposures during key periods – birth and early life, childhood, and early adulthood - to understand the stages of life where exposure is important.

In the following sections, we provide an overview of the relevant literature (Section 2), outline the data and methods used to address our research questions (Section 3), present the results (Section 4) and discuss the conclusions and implications of the findings (Section 5).

## 2. Literature review

### Birth and early life

Previous research has hypothesised that poor intrauterine growth, manifested as low birthweight, may lead to a decreased peak number of primordial follicles, which in turn may be associated with earlier menopause (Mishra et al 2019). Several studies investigated the relationship between low birth weight and early menopause (Bjelland et al 2020; Ruth et al. 2016; Steiner et al 2010; Tom et al 2010; Hardy and Kuh 2002; Treloar et al 2000; Cresswell et al 1997), but only few found a significant association (Bjelland et al 2020; Ruth et al. 2016; Steiner et al 2010; Tom at al. 2010). In one of the few studies which had information on gestational age, Tom et al. (2010) also illustrated an increased risk for menopause before 45 for women with heavy weight at birth and high birthweight standardized by gestational age - a marker of faster fetal growth rate – indicating that growth rate rather than prematurity might be more important. Some studies showed a significant association with early menopause of low weight at 1 year (Cresswell et al 1997) or at 2 years (Hardy and Kuh 2002).

Nutritional status may influence menopause, as children who are not breastfed, who have poor early growth, and who are malnourished tend to experience an earlier menopause (Ettorre and Bachmann 2019; Gold 2011). Breastfeeding was found to delay menopause for women in the 1946 National Survey of Health and Development (NSHD) (Mishra et al 2007; Hardy and Kuh 2002), but no association was found in Ruth et al.’s (2016) UK Biobank study.

It has been hypothesised that prenatal exposure to cigarette smoke may affect the follicle pool, formed during fetal development, by suppressing the formation of follicles or by damaging them (Tawflik et al 2015). A negative effect of maternal smoking around birth on early menopause was observed in UK Biobank (Ruth et al 2016), while other studies suggested that the effect of prenatal exposure to cigarette smoke may act via adult smoking (Strohsnitter et al 2008) or found no association (Ernst et al 2012; Hart et al 2009; Steiner et al 2010).

It is hypothesised that at both ends of the female reproductive span the risk of reproductive problems may be increased (Hardy and Kuh 2002) as if the maternal cycle is irregular at conception the maturational state of the oocyte at ovulation may not be optimal, which increases the risk of ovarian maldevelopment, and thus infertility and early menopause in the daughter (Smits et al 1999). However, Steiner et al (2010) showed that having a mother aged 35 years or older at birth was associated with later age at menopause compared with mothers aged 20–34 years.

### Socioeconomic position

Several studies have indicated an association between disadvantaged childhood socioeconomic position and earlier menopause, and the impact of socioeconomic disadvantage has been considered to act through the effects of hardship experience throughout life and health behaviours (Mishra et al 2009; Hardy and Kuh 2005; Lawlor et al 2003). Others have found no or weak associations (Kuh et al 2005; Cresswell et al 1997). Socioeconomic position in childhood has been found to be more strongly related to early menopause than adult socioeconomic position in the 1946 NSHD (Mishra et al 2009; Hardy and Kuh 2005), while adverse socioeconomic position from childhood through to adulthood was associated with a younger age at menopause in a sub-study of the British Women’s Heart and Health Study (Lawlor et al 2003). Adult social class was not related to early menopause in a study using 1958 NCDS data (up until age 45) (Tom et al 2010).

### Psychological distress

Psychological health in early life has been linked to the onset of menopause, potentially mediated through the function of the hypothalamic-pituitary-adrenal axis, which affects the reproduction function (Mishra et al 2007). For example, parental separation, hypothesised as an indicator of early emotional stress, has been associated with earlier menopause in the 1946 NSHD (Hardy and Kuh 2005).

### Cognitive ability

Several studies have found that poorer cognitive ability in childhood is associated with early menopause with earlier measures of cognitive ability being more strongly related to age at menopause than tests taken later in life (Mishra et al 2007; Kuh et al 2005; Whalley et al 2004). Brain development and reproductive aging may both be influenced by ovarian steroids operating from early life onwards, perhaps through a similar effect on neuron and oocyte loss (Hardy and Kuh 2002; Finch and Kirckwood 2000). Heritability studies suggested potential “genetic programming”, raising the prospect that there are genes associated with menopause that are also associated with neurocognitive function (van Asselt 2004; Trealor 1998).

### Reproductive health

It has been hypothesized that fewer ovulatory cycles, crudely estimated by late age at menarche, longer menstrual cycles, use of oral contraceptives, and a higher number of full-term pregnancies – events contributing to periods of anovulation - result in later age at menopause (Cramer et al 1995). The evidence, however, is contested (Mishra et al 2017; Ruth et al 2016; Gold 2011; Hardy and Kuh 1999; van Noord et al 1997; Torgerson 1994).

UK Biobank data showed no evidence of a significant association between gynaecological problems and early menopause (Ruth et al 2016). However, several studies investigating the impact of pelvic or ovarian infections, or routine gynaecological surgeries, on fertility and ovarian function, primarily by studying response to ovarian stimulation during IVF cycles, have shown that women who respond poorly to ovarian stimulation are likely to become menopausal earlier (Fenton 2012; Subrat et al 2012; Nikolaou and Templeton 2003; Nikolaou et al 2002; Lawson et al 2003).

### Health behaviour

Behavioural factors such as smoking, drinking and exercise affect the endocrine function and women’s reproductive hormones, and thus may have an impact on the timing of menopause.

Cigarette smoking has multiple effects on hormone secretion, mainly mediated by the pharmacological action of nicotine and by toxins such as thiocyanate (Kapoor and Jones 2005). It has been shown that women who smoke are more likely to experience menopause between 1 and 2 years earlier than non-smokers (Bjelland, et al 2018; Hayatbakhsh et al 2016; Schoenaker et al 2014; Gold 2011; Parente et al 2008). Pooled analysis of 17 studies demonstrated that higher intensity, longer duration, higher cumulative dose, earlier age at smoking initiation, and shorter time since quitting smoking, were associated with higher risk of early menopause (Zhu et al 2018).

Several studies have postulated that moderate consumption of alcohol delays menopause (Taneri et al 2016; Sapre et al 2014; Torgerson et al 2007) by inducing oestrogen production (Gill 2000; Muti et al 1998). Other studies have shown association with younger age at menopause (Choi et al 2017) or found no relationship (Cooper et al 2001).

Moderate physical activity has been associated with later age at natural menopause (Schoenaker et al 2014) in some studies, while others observed no association for either adolescence or adulthood physical activity (Zhao et al 2018).

Associations between BMI and obesity with ovarian reserve and menopause have been inconsistent across studies and the mechanisms underlying the associations unclear (Bjelland et al 2018; Zhu et al 2018; Hardy et al 2008; Gold et al 2001; Broomberg et al 1997; Beser et al 1994). However, many studies used a BMI measure in midlife only, whereas the effects of BMI may depend on the stage of menopause transition (Santoro et al 2011). Past research has also shown that change in weight rather than weight at a particular point impacts the timing of menopause (Szegda et al 2017).

In summary, the literature on non-genetic risk factors for earlier age at menopause paints a complex picture suggesting many factors may be at play. We contribute to this literature with a prospective study where we observe women at all life stages across two birth cohorts. This permits us to investigate the effect of multiple factors on early menopause before the age of 45 focusing on the life stage at which they are particularly harmful (i.e. when the exposure is associated with the highest risk) as it may shed light on the biological origins of menopause and guide the timing for interventions (if the observed associations reflect causal effects).

## 3. Research questions, data, and methods

This study investigates risk factors for early menopause at three distinct stages of life (birth, childhood, and early adulthood), respecting the time sequence of exposures. We address the following research questions:

1. What birth characteristics are associated with menopause before 45 years of age?
2. What childhood characteristics are associated with early menopause, while factors at birth are accounted for?
3. And lastly, what adult characteristics influence early menopause, while early life and childhood factors are accounted for?

The data used in this study is harmonized pooled individual level data comprising women from the 1958 National Child Development Study (NCDS) (See: http://www.cls.ucl.ac.uk/ncds) and the 1970 British Cohort Study (BCS70) (See: http://www.cls.ucl.ac.uk/bcs70). Each study follows the lives of around 7,000 women (about 17,000 people) born in England, Scotland and Wales in a single week of 1958 and 1970, respectively. Since birth, study members of the NCDS cohort (and/or their parents) have been interviewed 10 times at ages 7, 11, 16, 23, 33, 42, 44/45, 46, 50 and 55. We use information collected at birth and ages 7, 11, 16, 33, 42, 44/45 and 50. Since birth, study members of the BCS70 cohort (and/or their parents) have been interviewed 9 times at ages 5, 10, 16, 26, 30, 34, 38, 42 and 46. We use information collected at birth and ages 5, 10, 16, 30, 38, 42 and 46.

### 3.1 Potential risk factors

The choice of potential risk factors is guided by the literature reviewed above. Risk factors at birth include: father’s social class, maternal smoking during pregnancy, birthweight standardized for gestational age, maternal age, and breastfeeding. Childhood risk factors include: cognitive ability at age 10 (BCS70) or 11 (NCDS), age at menarche, and a set of health behaviour characteristics at age 16 - smoking, alcohol consumption, physical activity, BMI and psychological distress. Adulthood risk factors include: the same a set of health behaviours - smoking, alcohol consumption, physical activity, BMI and psychological distress - measured at age 30 (BCS70) or 33 (NCDS). Other adult risk factors include a set of reproductive characteristics such as experience of gynaecological problems (by age 30 in BCS70 or 33 in NCDS), use of contraceptives (by age 30 in BCS70 or 42 in NCDS), and live births (by age 38 in BCS70 or 42 in NCDS); as well as a measure of occupational social class at 38 (BCS70) or 42 (NCDS). When not used as exposures, we control for (some of) these factors as potential confounders (see 3.3 Statistical analysis). Table A1 in Supplementary material details the variables used in this pooled analysis, following data harmonisation.

### 3.2 Menopause status

Menopause status is determined using information collected at age 42 and 46 in BCS70 and age 44/45 and 50 in NCDS. Women in both cohorts were asked if they had menstrual periods in the past 12 months. In the presence of amenorrhea, they were asked if they had periods in the past 3 months. Women with no periods in the past 12 months were asked about their age and the month at their final menstrual period (FMP) and the reason for amenorrhea. All women were asked about changes in the regularity of their menstrual cycles in the last few years or before their FMP, about undergone hysterectomy or bilateral oophorectomy, and hormone replacement therapy (HRT) use, including the dates of surgery or initiation of HRT. The derivation of menopause status followed widely accepted classification (Harlow et al 2012; WHO 1996). Natural menopause was defined as at least 12 consecutive months of amenorrhea not induced by surgery or other medical treatment. Peri-menopausal women were those with 3–11 months of amenorrhea or whose periods became less regular in the absence of amenorrhea. Pre-menopausal women reported menstruation within the last 3 months. Those who have undergone hysterectomy or bilateral oophorectomy prior to their FMP or whose periods stopped for other obvious reasons (e.g. pregnancy, contraceptives, chemotherapy or radiotherapy), as well as women who started HRT prior to the FMP, were classified in separate categories and were excluded from this analysis.^2^ A binary outcome variable distinguishing women experiencing menopause before age 45 from those who experienced menopause at 45 or more years, who were peri- or pre-menopause was defined. Details about the derivation and classification of women into menopause status are provided in Supplementary material (see Table A2).

### 3.3 Statistical analysis

Descriptive statistics for all variables included in the analysis for each cohort separately, including means and standard deviation, median and range for continuous variables, and percentages with 95% confidence intervals for categorical variables, are presented in Table A3 in Supplementary material.

We use multivariable logistic regression and present adjusted odds ratios and 95% confidence intervals for the association between each potential risk factor and menopause before 45 years after controlling for confounders. The data for the two cohorts are pooled with models incorporating a dummy variable identifying which cohort the woman belonged to (1958 or 1970). We use multiple imputation techniques to tackle data missingness (see section 3.4). Respecting the temporal sequence of exposures, we fit a series of models to estimate associations of risk factors adjusted for preceding variables only. We estimate the association between early menopause and: social class at birth controlling for cohort year only; maternal smoking controlling for social class; birthweight (standardized for gestational age) controlling for social class, maternal smoking, and maternal age at birth; breastfeeding controlling for all other birth variables. We estimate the association between early menopause and cognition (at age 10 or 11) controlling for all birth factors. We then fit a model including all other childhood risk factors, controlling for cognitive ability and birth factors.^3^ Finally, a model was used to estimate the associations between early menopause and all adulthood risk factors, controlling for childhood and birth factors.^4^ We also fit models for each potential risk factor adjusted for cohort year only and the results are presented in Supplementary material (see table A4).

The pooled cohort analysis enables us to estimate associations with increased statistical power (due to the increased size of our analytical sample) and focus on the common factors affecting menopause before 45 (rather than associations observed in each study separately).

In secondary analyses, we did, however, carry out logistic regression analyses relating menopause before 45 to the birth, childhood and early adulthood variables, in each cohort separately. These results are presented in Supplementary material, tables A5 and A6. All analyses were conducted using Stata version 15.0.

### 3.4 Missing data

To avoid the limitation of performing this analysis on a significantly reduced analytical sample, due to simultaneous inclusion of a large number of variables from different periods of life and with different levels of unit and item missingness (missing data in risk factors is between 4-31% in NCDS and 3-57% in BCS70; see Table A3 in Supplementary material), we used Multiple Imputation (MI) with chained equations (Mostafa et al 2021; White et al. 2011; Von Hippel 2007), under an assumption of missing at random^5^, performed in each cohort separately. The imputation model included all potential risk factors (exposures) and menopause before 45 years (outcome), but missing values were only imputed on exposures (see details about menopause status missingness in Tables A2.1 and A2.2 in Supplementary material). Fifty imputed datasets were created for each cohort and models were subsequently fitted using the combined (NCDS and BCS) imputed datasets. The imputation was conducted using Stata version 15.0.

## 4. Results

The analytical sample comprised 6,805 (natural) menopausal, peri- and pre-menopausal women (3,191 participating in the 1970 and 3,614 participating in the 1958 cohort)^6^. By age 45, 6.5% of women in BCS70 and 10.2% of women in NCDS analytical samples had undergone natural menopause. The pooled sample comprised of 8.4% early menopausal women. Table A3 in Supplementary material shows women’s birth, childhood, and adult characteristics in each cohort separately.

### 4.1 Birth models

Table 4.1 reports associations between women’s traits at birth and the subsequent probability of early menopause. Women whose fathers at birth worked in manual occupations had a twofold increase in the odds for early menopause compared to women whose fathers occupied non-manual jobs; the increase in the odds is 2.2 times higher for women whose fathers were absent at the time of their birth. Women whose mothers smoked during pregnancy had 24% higher odds for early menopause compared to women whose mothers did not smoke during pregnancy. Women who were breastfed for less than a month had 30% higher odds for early menopause compared to women who were breastfed for one month or longer.

**Table 4.1.**
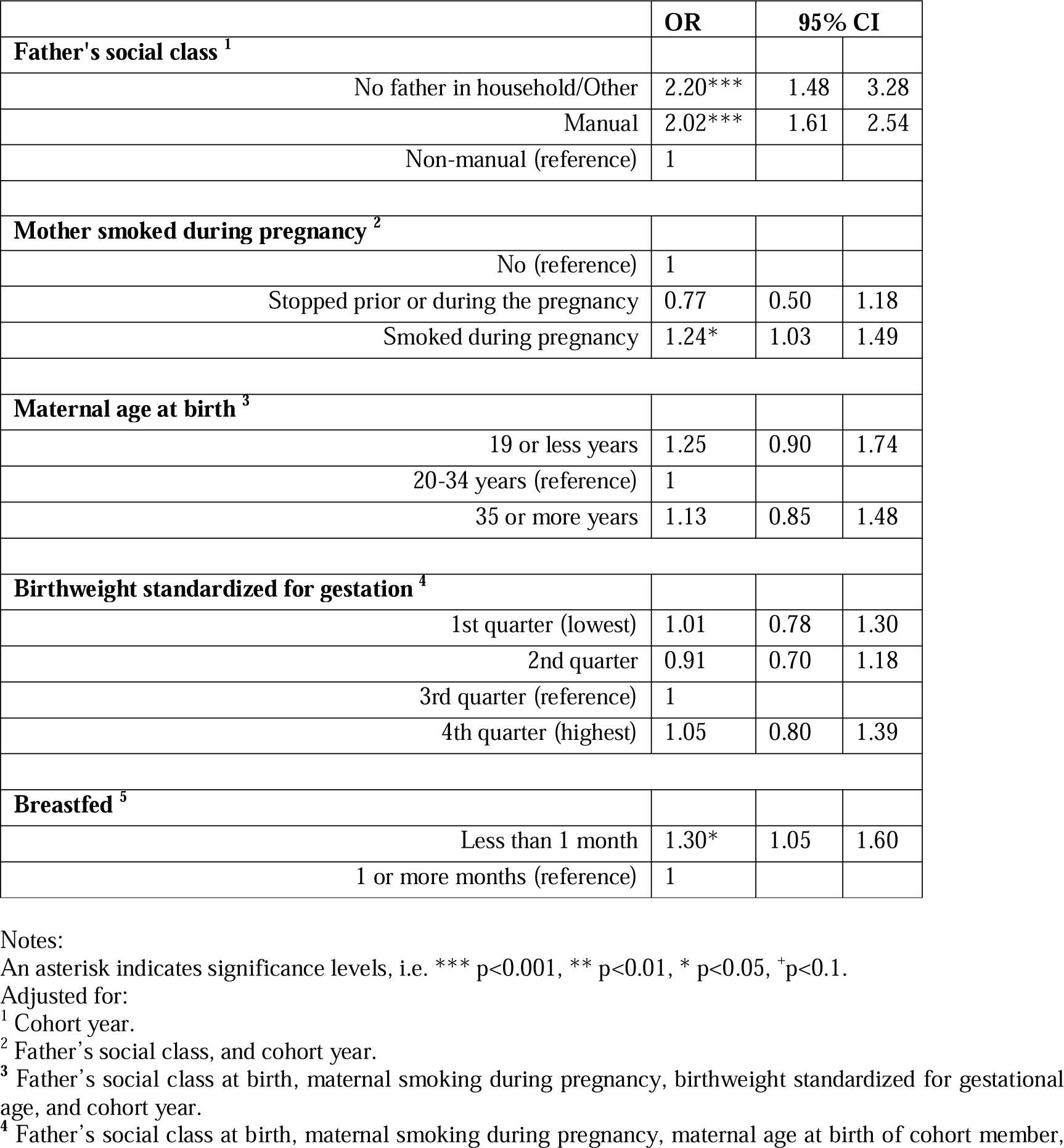
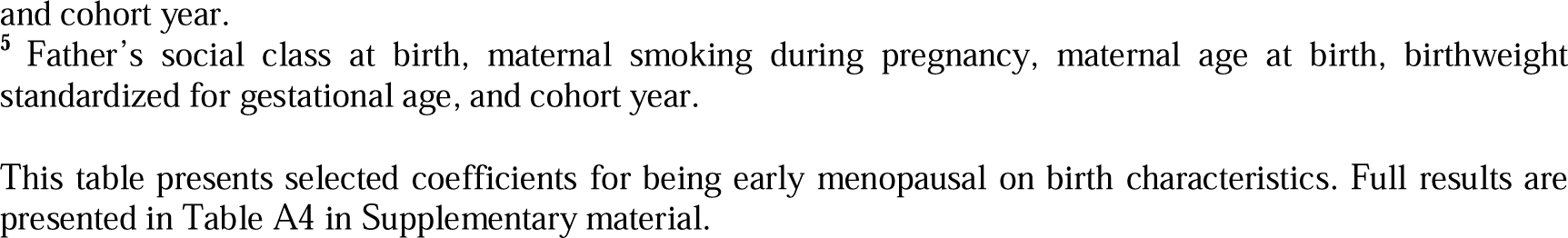
Odds Ratios (OR) and 95% confidence intervals (CI) for being early menopausal on birth characteristics (n=6,805)

|  | OR | 95% CI |  |
| --- | --- | --- | --- |
| **Father's social class <sup>1</sup>** |  |  |  |
| No father in household/Other | 2.20*** | 1.48 | 3.28 |
| Manual | 2.02*** | 1.61 | 2.54 |
| Non-manual (reference) | 1 |  |  |
| **Mother smoked during pregnancy <sup>2</sup>** |  |  |  |
| No (reference) | 1 |  |  |
| Stopped prior or during the pregnancy | 0.77 | 0.50 | 1.18 |
| Smoked during pregnancy | 1.24* | 1.03 | 1.49 |
| **Maternal age at birth <sup>3</sup>** |  |  |  |
| 19 or less years | 1.25 | 0.90 | 1.74 |
| 20-34 years (reference) | 1 |  |  |
| 35 or more years | 1.13 | 0.85 | 1.48 |
| **Birthweight standardized for gestation <sup>4</sup>** |  |  |  |
| 1st quarter (lowest) | 1.01 | 0.78 | 1.30 |
| 2nd quarter | 0.91 | 0.70 | 1.18 |
| 3rd quarter (reference) | 1 |  |  |
| 4th quarter (highest) | 1.05 | 0.80 | 1.39 |
| **Breastfed <sup>5</sup>** |  |  |  |
| Less than 1 month | 1.30* | 1.05 | 1.60 |
| 1 or more months (reference) | 1 |  |  |

Notes:
An asterisk indicates significance levels, i.e. \*\*\* p<0.001, \*\* p<0.01, \* p<0.05, +p<0.1.
Adjusted for:
<sup>1</sup> Cohort year.
<sup>2</sup> Father's social class, and cohort year.
<sup>3</sup> Father's social class at birth, maternal smoking during pregnancy, birthweight standardized for gestational age, and cohort year.
<sup>4</sup> Father's social class at birth, maternal smoking during pregnancy, maternal age at birth of cohort member,
and cohort year.
<sup>5</sup> Father's social class at birth, maternal smoking during pregnancy, maternal age at birth, birthweight standardized for gestational age, and cohort year.
This table presents selected coefficients for being early menopausal on birth characteristics. Full results are presented in Table A4 in Supplementary material.

Younger age of mother at birth was associated with increased odds, but the 95% confidence interval included 1. There was no evidence for association between birthweight (standardized for gestational age) and early menopause.

### 4.2 Childhood models

Table 4.2 presents associations between childhood characteristics and early menopause. The odds for early menopause decreased with an increase in childhood cognitive ability. One standard deviation higher cognitive ability score corresponded to 36% lower odds for being early menopausal. Women who smoked at 16, compared to those who did not, had 51% higher odds for early menopause.

**Table 4.2.** Odds Ratios (OR) and 95% confidence intervals (CI) for being early menopausal on childhood characteristics (n=6,805)

|  | OR | 95% CI |  |
| --- | --- | --- | --- |
| **Cognitive ability at 10/11 (per SD) <sup>1</sup>** | 0.64*** | 0.57 | 0.71 |
| **Age at menarche (per year)** | 0.98 | 0.90 | 1.07 |
| **BMI at 16 (per kg/m2)** | 1.03 | 0.99 | 1.07 |
| **Smoking at 16** |  |  |  |
| No (reference) | 1 |  |  |
| Yes | 1.51*** | 1.19 | 1.92 |
| **Drinking the week prior age 16 interview** |  |  |  |
| No (reference) | 1 |  |  |
| Yes | 1.09 | 0.85 | 1.39 |
| **Exercise at 16** |  |  |  |
| Monthly or less often (reference) | 1 |  |  |
| Weekly (Often) | 0.92 | 0.70 | 1.20 |
| **Emotional/Neurotic score at 16** | 1.00 | 0.92 | 1.10 |

Notes:
An asterisk indicates significance levels, i.e. \*\*\* p<0.001, \*\* p<0.01, \* p<0.05, +p<0.1.
<sup>1</sup> Adjusted for all birth characteristics, and cohort year.
The second childhood model (from which all the other coefficients come from) included all remaining potential risk factors in childhood (age at menarche, BMI, smoking, alcohol drinking, exercise, and emotional/neurotic score), adjusted for cognitive ability at 10/11 and
birth characteristics, and cohort year.
This table presents selected coefficients for being early menopausal on childhood characteristics. Full results are presented in Table A4 in Supplementary material.

There was no evidence that menarcheal age, alcohol drinking, exercise, or emotional problems at 16, were associated with menopause before 45.

### 4.3 Early adulthood model

Table 4.3 presents associations between characteristics recorded in women’s early adulthood, and their probability of early menopause. Women who suffered periods or other gynaecological problems by their early thirties, and those who smoked in their early thirties, were 68% and 69% more likely to be menopausal before 45, respectively. On the other hand, alcohol drinking, once to three times per month, compared to less frequent consumption, was associated with decreased odds for early cessation of menstrual periods, although there was no evidence of a trend across categories. Further, the odds for early menopause decreased with one or more days of exercise per week (as compared to less frequent exercise). Women who were not economically active in their late thirties or early forties were more likely to undergo early menopause.

**Table 4.3.**
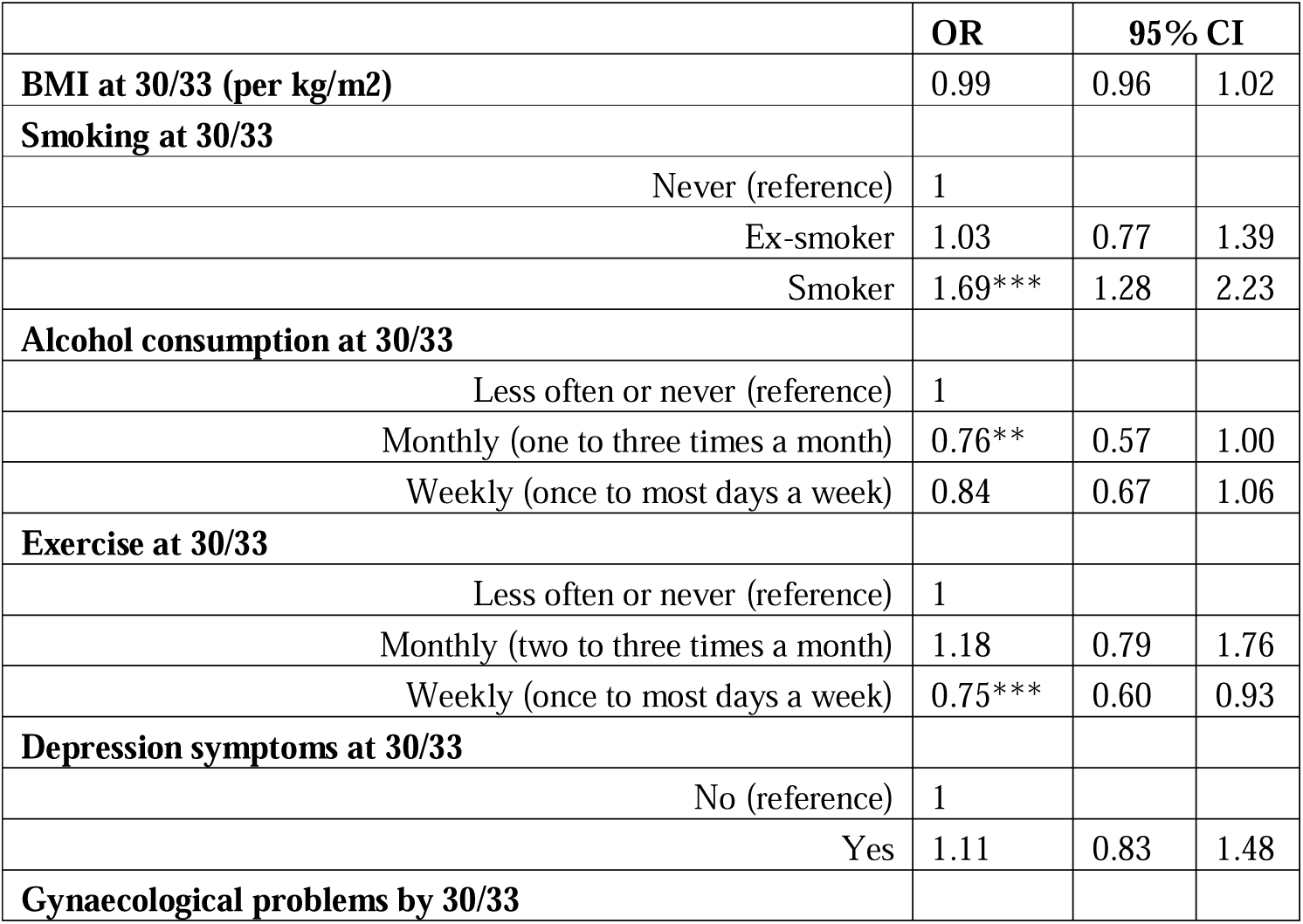
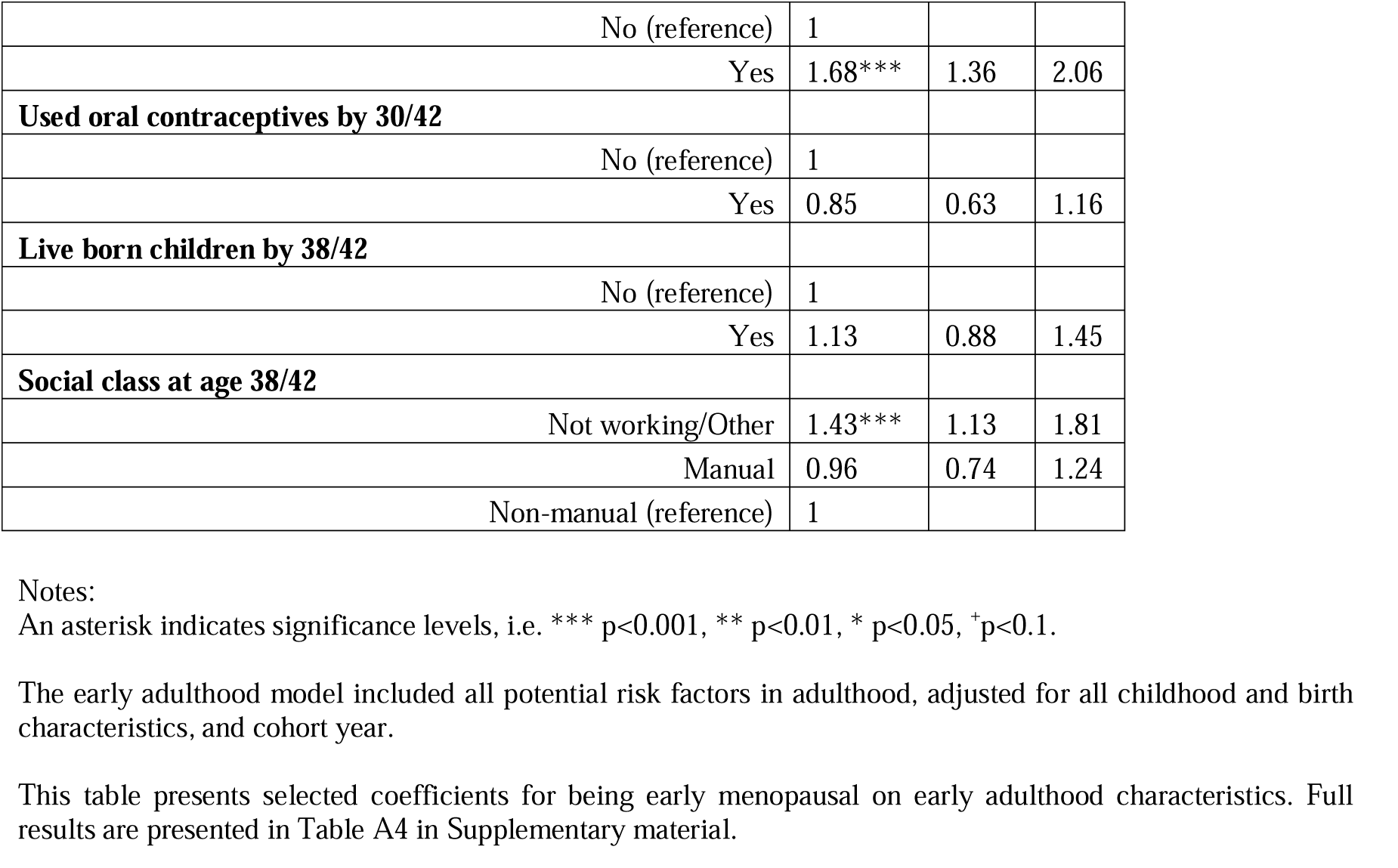
Odds Ratios (OR) and 95% confidence intervals (CI) for being early menopausal on early adulthood characteristics (n=6,805)

|  | **OR** | **95% CI** |  |
| --- | --- | --- | --- |
| **BMI at 30/33 (per kg/m2)** | 0.99 | 0.96 | 1.02 |
| **Smoking at 30/33** |  |  |  |
| Never (reference) | 1 |  |  |
| Ex-smoker | 1.03 | 0.77 | 1.39 |
| Smoker | 1.69*** | 1.28 | 2.23 |
| **Alcohol consumption at 30/33** |  |  |  |
| Less often or never (reference) | 1 |  |  |
| Monthly (one to three times a month) | 0.76** | 0.57 | 1.00 |
| Weekly (once to most days a week) | 0.84 | 0.67 | 1.06 |
| **Exercise at 30/33** |  |  |  |
| Less often or never (reference) | 1 |  |  |
| Monthly (two to three times a month) | 1.18 | 0.79 | 1.76 |
| Weekly (once to most days a week) | 0.75*** | 0.60 | 0.93 |
| **Depression symptoms at 30/33** |  |  |  |
| No (reference) | 1 |  |  |
| Yes | 1.11 | 0.83 | 1.48 |
| **Gynaecological problems by 30/33** |  |  |  |
| No (reference) | 1 |  |  |
| Yes | 1.68*** | 1.36 | 2.06 |
| **Used oral contraceptives by 30/42** |  |  |  |
| No (reference) | 1 |  |  |
| Yes | 0.85 | 0.63 | 1.16 |
| **Live born children by 38/42** |  |  |  |
| No (reference) | 1 |  |  |
| Yes | 1.13 | 0.88 | 1.45 |
| **Social class at age 38/42** |  |  |  |
| Not working/Other | 1.43*** | 1.13 | 1.81 |
| Manual | 0.96 | 0.74 | 1.24 |
| Non-manual (reference) | 1 |  |  |

Notes:
An asterisk indicates significance levels, i.e. \*\*\* $p < 0.001$ , \*\* $p < 0.01$ , \* $p < 0.05$ , + $p < 0.1$ .
The early adulthood model included all potential risk factors in adulthood, adjusted for all childhood and birth characteristics, and cohort year.
This table presents selected coefficients for being early menopausal on early adulthood characteristics. Full results are presented in Table A4 in Supplementary material.

Other factors, such as BMI, mental health status, use of oral contraceptives or nulliparity were not significantly associated with early menopause.

## 5. Discussion

We found that multiple factors from birth, childhood and adulthood are associated with menopause before 45 years of age. The early life factors associated with early menopause were father in a manual job, or no father figure at birth, mother smoking during pregnancy, and the absence of or short duration of breastfeeding. Childhood factors increasing the likelihood of early menopause were poor cognitive ability and smoking. Early adulthood factors associated with an increased probability of early menopause were smoking, no alcohol consumption, lower levels of exercise, gynaecological problems and not working.

Our findings support the hypothesis that fetal and early life experiences are associated with age at menopause. Consistent with previous studies, we find socioeconomic position, captured here by father’s social class at birth, to be related to the timing of menopause. The mechanism underlying this association is unclear but may act through health behaviour such as diet and nutrition or may be due to its relationship with early emotional stress, for example that caused by the absence of a parent or other adverse experiences as suggested by previous studies (Hardy and Kuh 2005; Lawlor et al 2003).

Our findings on the role of maternal smoking are consistent with the hypothesis that prenatal exposure to cigarette smoking may supress the formation of the ovarian follicle pool or damage it (Tawflik 2015). Our results on an association between breastfeeding and early menopause are consistent with those from the NSHD (Hardy and Kuh 2002), described previously with the role of nutrition (Ettorre and Bachmann 2019; Gold 2011; Mishra et al 2007). Inconsistencies with other studies showing no association (e.g. Ruth et al 2016) may be due retrospective reporting of breastfeeding in such studies or factors controlled for in the analyses (Jivraj et al 2017).

Our observation of a strong association with childhood cognitive ability is consistent with findings from other British birth cohort studies (Kuh et al 2005), where “genetic programming” has been suggested as an explanation for this link (van Asselt 2004; Trealor 1998).

Our study adds to the already consistent evidence that cigarette smoking is associated with early menopause. We also support previous findings that earlier initiation of smoking is linked to earlier menopause (Zhu et al 2018) as we find increased odds of early menopause with smoking at age 16 years.

Smoking was not the only health-related behaviour associated with the timing of menopause. Physical activity and alcohol consumption were associated with lower odds for being early menopausal. Our analysis suggests that it is high levels of activity, in a woman’s early thirties, one to several days of exercise a week, which is associated with lower odds for early menopause. Previous research on the role of physical activity on menopause is scarce and inconclusive, however it has been demonstrated that women’s reproductive hormones can be affected by physical factors (Arena et al 1995). We found that drinking, one to three times a month, in a woman’s early thirties, compared to less frequent drinking, lowered the odds for early menopause, but that there was no difference in odds for heavier drinkers. Although this observation is in line with a study using UK Biobank data (Ruth et al 2016), previous research on alcohol has not been consistent. Some studies have suggested that moderate alcohol consumption increases oestrogen levels in women (Gill 2000; Muti et al 1998). However, other studies have argued that health benefits of light to moderate alcohol use, but not heavier, are likely to be spurious and due to control group misclassification (e.g. some abstainers may be former dependent or heavy drinkers, who quit because of poor health, or individuals with long-term chronic illness) and residual confounding (Maggs and Staff 2017).

Gynaecological health was strongly linked to early menopause in our study. Previous evidence on the impact of gynaecological health on menopause timing is scarce. Gynaecological problems were not associated with early menopause in a study using UK Biobank data (Ruth et al 2016) but have been linked to it indirectly in studies on (in)fertility (Nikolaou et al 2002). As different gynaecological conditions may have different effects on menopause, exploring the role of particular gynaecological problems (e.g. sexually transmitted infections) is important to better understand the relationship with menopause timing, especially as some of these problems might be preventable.

Contrary to the hypothesis (e.g. Mishra et al. 2017) our results do not indicate an association between age of menarche or live born children with the timing of menopause (in either crude or adjusted models). Noting the wide range of factors our models accounted for (in both childhood and adulthood), this may suggest that menarche or parity effects observed previously might be due to residual confounding or recall error. Age at menarche in our analyses is reported by mothers when daughters were aged 16 rather than self-reported in adulthood (Cooper et al. 2007); and nulliparity derived using pregnancy histories through the study lifecycle.

And lastly, we observed women not in employment in their late thirties or early forties had greater odds of early menopause. Previous findings have not been consistent including a study using NCDS data (up until age 45) which found no effect on menopause before 45 years (Tom et al 2010). This, again, may be indicative of the importance of the timing of risk factor exposure. We note, that in our study this measure is not strictly preceding the menopause but is likely representative of earlier adult socioeconomic position.

There are limitations to consider in our study, such as potential biases resulting from unobserved confounding. Even though we account for a wide range of prospectively collected risk factors, our analysis focuses on characteristics previously highlighted in the literature on risk factors for premature or early menopause and does not account for factors discussed less frequently in the literature, e.g. the role of women’s physical health or maternal health (physical, mental, reproductive). Other limitations include the retrospective collection of information on menopause and potential for recall bias, although recall is not over long period. Previous research has indicated that menopause is reasonably well recalled, the recall error is expected to increase with the time after menopause (Hahn et al 1997). Also, our analysis investigates common risk factors for women born 12 years apart which may obscure cohort-specific relationships. We justified the use of a combined study with comparable study designs and measures but acknowledge the potential for bias in the harmonized data that may result from combining data from different studies (e.g. due to age and periods effects, or different level of sample attrition and missing data) which may in turn affect the generalizability of our findings. Nonetheless, it is reassuring that the associations presented here are consistent with those observed in each cohort separately. If statistical significance is not reached in a cohort, the estimated effects remain in the same direction (see study-specific analyses in Tables A5 and A6 in Supplementary Material). Further, our analysis did not consider that premature (menopause before 40 years of age) and early menopause (between 40 and 44 years) might be impacted by different (non-genetic) factors, while previous genetic research has questioned whether premature menopause is determined by the same genes as menopause that occurs between 40 and 44 years (De Bruin et al 2001). As with any other longitudinal survey, both cohorts have been affected by attrition. Fortunately, the retention rates over the decades following the study members have been strong (Mostafa et al 2021). We approached the missing data problem using multiple imputation, but bias due to selective attrition cannot be ruled out.

In conclusion, our study demonstrated that characteristics at different stages of life are associated with early menopause – an event with serious health consequences affecting around one in ten women. Some of these associations may reflect genetic or underlying biological processes which, with further investigation, could lead to better understanding of the menopausal process and reproductive health across the life course. Further, some of the characteristics associated with early menopause relate to behaviours which are modifiable (e.g. breastfeeding, smoking, physical activity). If the presented associations reflect causal effects, early (timely) prevention may reduce the risks of early menopause and hence the adverse health outcomes associated with it.

## Data Availability

The data used in this study have been managed by the Centre for Longitudinal Studies - UCL and are accessible via the UK data archive (https://ukdataservice.ac.uk/).

## Acknowledgements

This project was funded by a grant from the Health Foundation [grant no. 546608].

## Footnotes

1 Formally, early menopause is defined as menopause between the ages of 40 and 44, and the onset of menopause before age 40 as premature menopause.

2 The same rules applied for the initial and follow-up sweep. However, once a woman had gone through a natural menopause or a surgery (hysterectomy/bilateral oophorectomy) or had initiated HRT prior to the FMP, their menopausal status remained throughout for the subsequent survey.

3 Respecting the temporal sequence of childhood risk factors, the association between age at menarche and early menopause has been considered without other age 16 variables (i.e. excluding BMI, smoking, drinking, exercise, and emotional distress), controlling for cognitive ability and factors at birth. No difference from the presented estimate was apparent (results not shown).

5 It implies that systematic differences between the missing and observed values can be explained by the observed data.

6 It excluded twins and triplets and women with missing multiple birth information, retaining only singletons. This is because previous research has established strong genetics relationship between being a child of a multiple birth and premature or early menopause (Mishra et al 2019).

## References

Anagnostis, P., Christou, K., Artzouchaltzi, A.M., Gkekas, N.K., Kosmidou, N., Siolos, P., et al., 2019. Early menopause and premature ovarian insufficiency are associated with increased risk of type 2 diabetes: a systematic review and meta-analysis, Eur. J. Endocrinol. 180, 41–50. https://doi.org/10.1530/EJE-18-0602.

Arena, B., Maffulli, N., Maffulli, F. et al., 1995. Reproductive Hormones and Menstrual Changes with Exercise in Female Athletes. Sports Med. 19, 278–287. https://doi.org/10.2165/00007256-199519040-00005.

Baron, J.A., La Vecchia, C., Levi, F., 1990. The antiestrogenic effect of cigarette smoking in women. Am J Obstet Gynecol. 167:502–514. https://doi.org/10.1016/0002-9378(90)90420-C.

Beser, E., Aydemir, V., Bozkaya, H., 1994. Body mass index and age at natural menopause. Gynecol Obstet Invest. 37, 40–2. https://doi.org/10.1159/000292518.

Bjelland, E.K., Gran, J.M., Hofvind, S., Eskild, A., 2020. The association of birthweight with age at natural menopause: a population study of women in Norway. Int J Epidemiol. 49, 528–536. https://doi.org/10.1093/ije/dyz207.

Bjelland, E.K., Hofvind, S., Byberg, L., Eskild, A., 2018. The relation of age at menarche with age at natural menopause: a population study of 336 788 women in Norway. Hum.Reprod. 33, 1149–1157. https://doi.org/10.1093/humrep/dey078.

Broomberg, J.T., Matthews, K.A., Kuller, L.H., Wing, R.R., Meilahn, E.N., Plantinga, P., 1997. Prospective study of the determinants of age at menopause. Am. J. Epidemiol. 145, 124–133. https://doi.org/10.1093/oxfordjournals.aje.a009083.

Bruin, J., Bovenhuis, H., Noord, P.A.H., Pearson, P., Arendonk, J., te Velde, E., Kuurman, W., Dorland, M., 2001. The Role of Genetic Factors in Age at Natural Menopause. Hum.Reprod. 16, 2014–8. https://doi.org/10.1097/00006254-200204000-00020.

Choi, J.I., Han, K., Lee, D.W., Kim, M.J., Shin, Y.J., Lee, H.N., 2017. Relationship between alcohol consumption and age at menopause: The Korea National Health and Nutrition Examination Survey, Taiwanese Journal of Obstetrics and Gynaecology 56, 482–486, https://doi.org/10.1016/j.tjog.2017.05.002.

Cooper, G.S., Baird, D.D., Darden, F.R., 2001. Measures of Menopausal Status in Relation to Demographic, Reproductive, and Behavioral Characteristics in a Population-based Study of Women Aged 35–49 Years. Am J Epidemiol. 153, 1159–1165. https://doi.org/10.1093/aje/153.12.1159.

Cooper, R., Blell, M., Hardy, R., et al., 2006. Validity of age at menarche self-reported in adulthood. Journal of Epidemiology & Community Health 60, 993–997.

Cramer, D.W., Xu, H., Harlow, B. L., 1995. Family history as a predictor of early menopause. Fertil. Steril. 64, 740–745. https://doi.org/10.1016/S0015-0282(16)57849-2.

Cresswell, J.L., Egger, P., Fall, C.H.D., Osmond, C., Fraser, R.B., Barker, D.J.P., 1997. Is the age of menopause determined in-utero? Early Hum. Dev. 49, 143–148.

Ernst, A., Kristensen, S.L., Toft, G.A., Thulstrup, M.L., Håkonsen, B., Olsen, S.F., Ramlau-Hansen, C.H., 2012. Maternal smoking during pregnancy and reproductive health of daughters: a follow-up study spanning two decades. Hum.Reprod. 27, 3593–3600. https://doi.org/10.1093/humrep/des337.

Ettorre, V.M., Bachmann, G.A., 2019. Childhood predictors of age at natural menopause, Case Reports in Women’s Health, 24, 2019, https://doi.org/10.1016/j.crwh.2019.e00148.

Fenton, A., Panay, N., 2012. Does routine gynecological surgery contribute to an early menopause? Climacteric 15, 1–2. https://doi.org/10.3109/13697137.2012.647623.

Finch, C.E., Kirkwood, T.B.L., 2000. Chance, Development and Aging. Oxford University Press, Oxford.

Gallagher, J.C., 2007. Effect of early menopause on bone mineral density and fractures. Menopause 14, 567–571. https://doi.org/10.1097/gme.0b013e31804c793d.

Gill, J. 2000. THE EFFECTS OF MODERATE ALCOHOL CONSUMPTION ON FEMALE HORMONE LEVELS AND REPRODUCTIVE FUNCTION. Alcohol and Alcoholism 35, 417–423. https://doi.org/10.1093/alcalc/35.5.417.

Gold, E.B., 2011. The Timing of the Age at Which Natural Menopause Occurs. Obstet Gynecol Clin North Am. 38, 425–440. https://doi.org/10.1016/j.ogc.2011.05.002.

Gold, E. B., Bromberger, J., Crawford, S., Samuels, S., Greendale, G. A., Harlow, S. D., Skurnick, J., 2001. Factors associated with age at natural menopause in a multiethnic sample of midlife women. Am. J. Epidemiol. 153, 865–873. https://doi.org/10.1093/aje/153.9.865.

Hardy, R., Kuh, D., 1999. Reproductive characteristics and age at inception of the perimenopause in a British national cohort. Am. J. Epidemiol. 149, 612–620. https://doi.org/10.1093/oxfordjournals.aje.a009861.

Hardy, R., Kuh, D., 2002. Does early growth influence timing of the menopause? Evidence from a British birth cohort. Hum.Reprod. 17, 2474–2479. https://doi.org/10.1093/humrep/17.9.2474.

Hardy, R., Kuh, D., 2005. Social and environmental conditions across the life course and age at menopause in a British birth cohort study. Int. J. Obs. and Gynae. 112, 346–354. https://doi.org/10.1111/j.1471-0528.2004.00348.x.

Hardy, R., Mishra, G.D., Kuh, D., 2008. Body mass index trajectories and age at menopause in a British birth cohort. Maturitas 59, 304–14. https://doi.org/10.1016/j.maturitas.2008.02.009.

Harlow, S.D, Gass, M., Hall, J.E., Lobo, R., Maki, P., Rebar, R.W., Sherman, S., Sluss, P.M., de Villiers, T.J., 2012. STRAW 10 Collaborative Group. Executive summary of the Stages of Reproductive Aging Workshop + 10: addressing the unfinished agenda of staging reproductive aging. Menopause 19, 387–95. https://doi.org/10.1097/gme.0b013e31824d8f40.

Hart, R., Sloboda, D.M., Doherty, D.A., Norman, R.J., Atkinson, H.C., Newnham, J.P., Dickinson, J.E., Hickey, M., 2009. Prenatal Determinants of Uterine Volume and Ovarian Reserve in Adolescence. J. Clin Endocrinol Metab. 94, 4931–4937. https://doi.org/10.1210/jc.2009-1342.

Hayatbakhsh, M.R., Clavarino, A., Williams, G.M., Sina, M., Najman, J.M., 2012. Cigarette smoking and age of menopause: a large prospective study. Maturitas 72, 346–52. https://doi.org/10.1016/j.maturitas.2012.05.004.

Hahn, R. A., Eaker, E., Rolka, H. 1997. Reliability of reported age at menopause. Am J Epidemiol 146, 771–775.

Jacob, C., Baird, J., Barker, M., Cooper, C., Hanson, M., 2017. The Importance of a Life Course Approach to Health: Chronic Disease Risk from Preconception through Adolescence and Adulthood: White paper World Health Organisation. https://eprints.soton.ac.uk/436656/

Jivraj, S., Goodman, A., Ploubidis, G. B., de Oliveira, C., 2017. Testing Comparability Between Retrospective Life History Data and Prospective Birth Cohort Study Data. J Gerontol B Psychol Sci Soc Sci. 75, 207–217. https://doi.org/10.1093/geronb/gbx042.

Kapoor, D., Jones, T. H., 2005. Smoking and hormones in health and endocrine disorders. Eur J Endocrinol. 152, 491–499. https://doi.org/10.1530/eje.1.01867

Kok, H.S, van Asselt, K.M., Schouw van der, Y.T., Peeters, P.H., Wijmenga, C., 2005. Genetic studies to identify genes underlying menopausal age. Hum. Reprod. Update. 11, 483–493. https://doi.org/10.1093/humupd/dmi024.

Kok, H. S., Kuh, D., Cooper, R., van der Schouw, Y. T., Grobbee, D. E., Wadsworth, M. E., Richards, M., 2006. Cognitive function across the life course and the menopausal transition in a British birth cohort. Menopause 13, 19–27. https://doi.org/10.1097/01.gme.0000196592.36711.a0.

Kuh, D., Butterworth, S., Kok, H., Richards, M., Hardy, R., Wadsworth, M., Leon, D., 2005. A Childhood cognitive ability and age at menopause: evidence from two cohort studies. Menopause 12, 475–482. https://doi.org/10.1097/01.GME.0000153889.40119.4C.

Lawlor, D.A., Ebrahim, S., Smith, G.D., 2003. The association of socio-economic position across the life course and age at menopause: the British Women’s Heart and Health Study. Int. J. Obs. and Gynae. 110, 1078–1087. https://doi.org/10.1111/j.1471-0528.2003.02519.x.

Whalley, L.J., Fox, H.C., Starr, J.M., Deary, I.J., 2004. Age at natural menopause and cognition. Maturitas 49, 148–156. https://doi.org/10.1016/j.maturitas.2003.12.014

Lawson, R., El-Toukhy, T., Kassab, A., Taylor, A., Braude, P., Parsons, J., Seed, P., 2003. Poor response to ovulation induction is a stronger predictor of early menopause than elevated basal FSH: a life table analysis. Hum. Reprod. 18, 527–533. https://doi.org/10.1093/humrep/deg101.

Maggs, J.L., Staff, J., 2017. No Benefit of Light to Moderate Drinking for Mortality From Coronary Heart Disease When Better Comparison Groups and Controls Included: A Commentary on Zhao et al. (2017). Journal of Studies on Alcohol and Drugs 78, 387–388. https://doi.org/10.15288/jsad.2017.78.387.

Mishra, G.D., Chung, H., Canob, A., Chedrauic, P., Goulisd, D.G., Lopese, P., Mueckg, A., Reesh, M., Levent, M., Senturki, M., Simoncinij, T., Stevensonk, J.C., Stutel, P., Tuomikoskim, P., Lambrinoudakin, I., 2019. Predictors of early menopause. Maturitas 123, 82–88. https://doi.org/10.1016/j.maturitas.2019.03.008.

Mishra, G.D., Pandeya, N., Dobson, A.J., Chung, H.F., Anderson, D., Kuh, D., Sandin, S., Giles, G.G., Bruinsma, F., Hayashi, K., Lee, J.S., Mizunuma, H., Cade, J.E., Burley, V., Greenwood, D.C., Goodman, A., Simonsen, M.K., Adami, H.O., Demakakos, P., Weiderpass, E., 2017. Early menarche, nulliparity and the risk for premature and early natural menopause. Hum. Reprod. 32, 679–686. https://doi.org/10.1093/humrep/dew350.

Mishra, G.D, Anderson, D., Schoenaker, D.A, Adami H.O, Avis, N.E, Brown, D., Bruinsma, F., Brunner, E., Cade, J.E., Crawford, S.L., Dobson, A. J., Elliott, J., Giles, G. G., Gold, E.B., Hayashi, K., Kuh, D., Lee, K.A., Lee, J.S., Melby, M.K., … Weiderpass, E., 2013. InterLACE: A new international collaboration for a life course approach to women’s reproductive health and chronic disease events. Maturitas 74, 235–240. https://doi.org/10.1016/j.maturitas.2012.12.011.

Mishra, G.D., Cooper, R., Tom, S.E., Kuh, D., 2009. Early life circumstances and their impact on menarche and Menopause. Women’s Health. 5, 175–190. https://doi.org/10.2217/17455057.5.2.175.

Mishra, G.D., Goodman, A., Koupil, I., 2007. Associations of birth characteristics with perimenopausal disorders: a prospective cohort study. Journal of Developmental Origins of Health and Disease. 10, 246–252. doi:10.1017/S204017441800065X.

Mostafa, T., Narayanan, M., Pongiglione, B., Dodgeon, B., Goodman, A., Silverwood, R.J., Ploubidis, G.B., 2021. Missing at random assumption made more plausible: evidence from the 1958 British birth cohort. J Clin Epidemiol, 136, 44–54. https://doi.org/10.1016/j.jclinepi.2021.02.019.

Muti, P., Trevisan, M., Micheli, A., Krogh, V., Bolelli, G., Sciaryno, R., Schunemann, H. J., Berrino, F., 1998. Alcohol consumption and total oestradiol in premenopausal women. Cancer Epidemiology, Biomarkers and Prevention 7, 189–193.

Nikolaou, D., Lavery, S., Turner, C., Margara, R., Trew, G., 2002. Is there a link between an extremely poor response to ovarian hyperstimulation and early ovarian failure? Hum. Reprod. 17, 1106–1111. https://doi.org/10.1093/humrep/17.4.1106.

Nikolaou, D., Templeton, A., 2003. Early ovarian ageing: a hypothesis Detection and clinical relevance. Hum Reprod 18, 1137–1139. https://doi.org/10.1093/humrep/deg245

Parente, R.C., Faerstein, E., Keller, R.C., Werneck, G.L., 2008. The relationship between smoking and age at the menopause: A systematic review. Maturitas 61, 287–298. https://doi.org/10.1016/j.maturitas.2008.09.021.

Ruth, K., Perry, J., Henley, W. et al., 2016. Events in Early Life are Associated with Female Reproductive Ageing: A UK Biobank Study. Sci Rep 6, 24710. https://doi.org/10.1038/srep24710.

Santoro, N., Randolph, J.F., 2011. Reproductive hormones and the menopause transition. Obstetrics and Gynecology Clinics of North America. 38, 455–466. https://doi.org/10.1016/j.ogc.2011.05.004.

Sapre, S., Thakur, R., 2014. Lifestyle and dietary factors determine age at natural menopause. J Mid-life Health 5, 3–5. https://www.jmidlifehealth.org/text.asp?2014/5/1/3/127779.

Schoenaker, D.A., Jackson, C.A., Rowlands, J.V., Mishra, G.D., 2014. Socioeconomic position, lifestyle factors and age at natural menopause: a systematic review and meta-analyses of studies across six continents. Int J Epidemiol. 43, 1542–62. https://doi.org/10.1093/ije/dyu094.

Shifren, J.L., Gass, M.L.S., 2014. The North American Menopause Society Recommendations for Clinical Care of Midlife Women. Menopause. 21, 1038–1062. https://doi.org/10.1097/gme.0000000000000319.

Smits, L.J., Zielhuis, G.A., Jongbloet, P.H., Bouchard, G., 1999. The association of birth interval, maternal age and season of birth with fertility of daughters: a retrospective cohort based on family reconstructions from nineteenth and early twentieth century Quebec. Paediatr Perinat Epidemiol. 13, 408–20.

Steiner, A.Z., D’Aloisio, A., DeRoo, L.A., Sandler, D.P., Baird, D.D., 2010. Association of Intrauterine and Early-Life Exposures With Age at Menopause in the Sister Study. Am. J. Epidemiol. 172. https://doi.org/10.1093/aje/kwq092.

Strohsnitter, W.C., Hatch, E.E., Hyer, M., Troisi, R., Kaufman, R.H., Robboy, S.J., Palmer, J.R., Titus-Ernstoff, L., Anderson, D., Hoover, R.N., Noller, K.L., 2008. The association between in utero cigarette smoke exposure and age at menopause. Am J Epidemiol. 167, 727–733. https://doi.org/10.1093/aje/kwm351.

Subrat, P., Santa, S.A., Vandana, J., 2012. The Concepts and Consequences of Early Ovarian Ageing: A Caveat to Women’s Health. J Reprod Infertil. 14, 3–7.

Szegda, K.L., Whitcomb, B.W., Purdue-Smithe, A.C., Boutot, M.E., Manson, J.E., Hankinson, S.E., Rosner, B.A, Bertone-Johnson, E.R., 2017. Adult adiposity and risk of early menopause. Hum Reprod. 32, 2522–2531. https://doi.org/10.1093/humrep/dex304.

Taneri, P.E., Kiefte-de Jong, J.C., Bramer, W.M., Daan, N.M, Franco, O.H, Muka, T., 2016 Association of alcohol consumption with the onset of natural menopause: a systematic review and meta-analysis. Hum. Reprod. Update. 22, 516–28. https://doi.org/10.1093/humupd/dmw013.

Tawfik, H., Kline, J., Jacobson, J., Tehranifar, P., Protacio, A., Flom, J.D., Cirillo, P., Cohn, B.A., Terry, M.B., 2015. Life Course Exposure to Smoke and Early Menopause and Menopausal Transition. Menopause 22, 1076–1083. https://doi.org/10.1097/GME.0000000000000444.

Tom S. E., Cooper, R., Kuh, D., Guralnik, J.M., Hardy, R., Power, C., 2010. Fetal environment and early age at natural menopause in a British birth cohort study. Hum. Reprod. 25, 791–798. https://doi.org/10.1093/humrep/dep451.

Torgerson, D.J., Avenellb, A., Russell, I.T., Reid, D.M., 1994. Factors associated with onset of menopause in women aged 45-49. Maturitas 19, 83–92. https://doi.org/10.1016/0378-5122(94)90057-4.

Torgerson, D.J., Thomas, R.E., Campbell, M.K., Reid, D.M., 1997. Alcohol consumption and age of maternal menopause are associated with menopause onset. Maturitas 26, 21–25.

Treloar, S.A., Do, K.A., Martin, N.G., 1998. Genetic influences on the age at menopause. Lancet 352, 1084–1085. https://doi.org/10.1016/S0140-6736(05)79753-1.

Treloar, S.A., Sadrzadeh, S., Do, K.A, Martin, N.G., Lambalk, C.B., 2000. Birth weight and age at menopause in Australian female twin pairs: exploration of the fetal origin hypothesis. Hum. Reprod. 15, 55–59.

van Asselt, K.M., Kok, H.S., Putter, H., Wijmenga, C., Peeters, P.H., Schouw van der, Y.T, Grobbee, D.E., te Velde, E.R., Mosselman, S., Pearson, P.L., 2004. Linkage analysis of extremely discordant and concordant sibling pairs identifies quantitative trait loci influencing variation in human menopausal age. Am J Hum Genet. 74, 444–453. https://doi.org/10.1086/382136.

van Noord, P.A.H., Dubas, S., Dorland, M., Boersma, H., te Velde, E., 1997. Age at natural menopause in a population-based screening cohort: the role of menarche, fecundity, and lifestyle factors. Fertil.Steril. 68, 95–102. https://doi.org/10.1016/s0015-0282(97)81482-3.

Von Hippel, P., 2007. Regression with missing Ys: an improved strategy for analyzing multiply imputed data. Sociological Methodology 37, 83–117.

White, I.R., Royston, P., Wood, A.M., 2011. Multiple imputation using chained equations: issues and guidance for practice. Statistics in Medicine 30, 377–399. https://doi.org/10.1002/sim.4067.

World Health Organization, 1996. Research on the menopause in the 1990s (Report of a WHO scientific group, WHO Technical Report Series, 886). Geneva World Health Organization.

Zhao, M., Whitcomb, B.W, Purdue-Smithe, A.C., Manson, J.E., Hankinson, S.E., Rosner, B.A., Bertone-Johnson, E.R., 2018. Physical activity is not related to risk of early menopause in a large prospective study. Hum Reprod. 33, 1960–1967. https://doi.org/10.1093/humrep/dey267.

Zhu, D., Chung, H.F., Dobson, A.J., Pandeya, N., Brunner, E.J., Kuh, D., Greenwood, D.C., Hardy, R., Cade, J.E., Giles, G.G., Bruinsma, F., Demakakos, P., Simonsen, M.K., Sandin, S., Weiderpass, E., Mishra, G.D. 2020. Type of menopause, age of menopause and variations in the risk of incident cardiovascular disease: pooled analysis of individual data from 10 international studies. Hum Reprod. 35, 1933–1943. doi: 10.1093/humrep/deaa124. PMID: 32563191.

Zhu, D., Chung, H.F., Dobson, A.J., Pandeya, N., Giles, G.G., Bruinsma, F., Brunner, E.J., Kuh, D., Hardy, R., Avis, N.E, Gold, E.B., Derby, C.A., Matthews, K.A., Cade, J.E., Greenwood, D.C., Demakakos, P., Brown, D.E., Sievert, L.L., Anderson, D., Hayashi, K., Lee, J.S., Mizunuma, H., Tillin, T., Simonsen, M.K., Adami, H.O., Weiderpass, E. Mishra, G.D., 2020. Age at natural menopause and risk of incident cardiovascular disease: a pooled analysis of individual patient data. Lancet Public Health. 4, 553–564. https://doi.org/10.1016/S2468-2667(19)30155-0.

Zhu, D., Chung, H.F., Pandeya, N., Dobson, A.J, Cade, J.E., Greenwood, D.C. et al., 2018. Relationships between intensity, duration, cumulative dose, and timing of smoking with age at menopause: A pooled analysis of individual data from 17 observational studies. PLoS Med. 15: e1002704. https://doi.org/10.1371/journal.pmed.1002704.

Zhu, D., Chung, H.F., Pandeya, N. et al., 2018. Body mass index and age at natural menopause: an international pooled analysis of 11 prospective studies. Eur J Epidemiol 33, 699–710. https://doi.org/10.1007/s10654-018-0367-y.

