## Supplementary material for "RISK FACTORS FOR EARLY NATURAL MENOPAUSE: EVIDENCE FROM THE 1958 AND 1970 BRITISH BIRTH COHORTS"

**Table A1. Questions and response options in 1970 BCS and 1958 NCDS cohorts and new variables (used in pooled analyses) created following data harmonisation**

| **1970 BCS cohort** | | | **1958 NCDS cohort** | | | **Harmonised** | | |
| --- | --- | --- | --- | --- | --- | --- | --- | --- |
| **Age (Respondent)** | **Question** | **Categories or scale** | **Age (Respondent)** | **Question** | **Categories or scale** | **Variable** | **Categories** | Note |
| Birth (midwife) | Birthweight of baby  Gestatational age | …lbs. …ozs. or …gm  ... days/…months | Birth (midwife) | Birth weight  Gestation period | …lbs. …ozs.  …days/…months | Birthweight (kg) standardized  by gestational age (weeks) | 1st quarter (lowest) 2nd quarter  3rd quarter  4th quarter (highest) | Birthweight was standardized by gestational age as follows: the difference between the woman’s birthweight and the mean birthweight was calculated for that gestational age and divided by the standard deviation of birthweight for that gestational age (Leon et al., 1998; Lawlor et al., 2005; Tom et al 2010). Birthweight for gestation z-scores was categorized into quarters. |
| Birth (mother) | Mother's age at birth | … years | Birth (mother) | Mother's age at birth | … years | Mother's age at birth | 19 or less years 20-34 years  35 or more years |  |
| Birth (mother) | Does the mother smoke now? If no: Did she ever smoke?  If yes: How long ago did she stop? How much does.did she smoke? Has she smoked during this  pregnancy? | Yes/No  Yes/No  Years/Months  1-4/5-14/15-24/25 or more/Not known  Yes/No/Not known | Birth (mother) | Did the patient smoke as many as  one cigarette a day during the 12 months before the start of this pregnancy? If so, how many per day during that period?  If smoked one per day  Did the patient change her smoking habits during this pregnancy? Record changes in the table below and month of pregnancy change made. | Did not smoke as many as 1 perd day Number smoked per day in that period  No change  Gave up/Month of pregnancy/Number per day smoked after change  Cut down/Month of pregnancy/Number per day  smoked after change  Increased/Month of pregnancy/Number per day  smoked after change | Has mother smoked during pregnancy | No  Stopped prior or during the pregnancy  Smoked during the pregnancy |  |
| Birth (mother) | Occupation of husband If self-employed  If not self-employed Employed at present Unemployed at present | Actual job, Description of job  Employing 25 or more persons/Employing fewer than 25 persons  Supervising others/Not supervising others | Birth (mother) | What was the husband's occupation?  If self-employed: Does he employ 10 or more persons?  If not self-employed: Does he supervise others (e.g. foreman, manager, charge- hand)? | Actual job / Industry  Yes/No  Yes/No | Father's social class at birth | Non-manual  Manual  No father in household | Categorized into “Non-manual”, including Registrar-General's Social class I, II and III: Professional occupations, Managerial and Other Professionals, and Non-manual skilled occupations, “Manual” - including Social class IV and V Semi-skilled and Unskilled workers, and “No father in household”. |
| Birth (mother) | Was {cohort member} breastfed partly  or wholly even for a few days? | Yes - for less than 1 month  Yes - for 1 month or more but less than 3 months  Yes - for 3 months or more  Yes but cannot remember for how long No, was not breastfed at all  Not known | Birth (mother) | Was the child breastfed (partly or wholly) as  a baby? | No  Yes - under 1 month Yes - over 1 month Don't know | Has been breastfed | Less than 1 month or never  Breastfed 1 or more months |  |
| 10 | Friendly Maths Test: A multiple choice test including arithmetic, number skills, fractions, algebra, geometry and statistics.  The Shortened Edinburgh Reading Test Godfrey Thompson Unit 1978): A test of word recognition, which examined vocabulary, syntax, sequencing, comprehension and retention. | Test scores ranged between 1-72 (included  72 items)  Test scores ranged between 0-67 (included  67 items) | 11 | Arithmetic/Mathematics Test: involving numerical and geometric work.  Reading Comprehension Test: involving selection of words which appropriately completed sentences | Test scores ranged between 1-40 (included 40 items)  Test scores ranged between 0-35 (included 35  items) | Cognititve ability score at 10/11 |  | Single main component (regression factor) score for cognition (using maths and reading tests at age 10 in BCS and age 11 in NCDS) extracted using Principal Components Analysis (PCA).  Source: Parsons, S. 2014. Childhood cognition in the 1970 British  Cohort Study: Data note. Centre for Longitudinal Studies, November 2014. https://cls.ucl.ac.uk/wp- content/uploads/2017/07/BCS70-Childhood- cognition-in-the-1970-British-Cohort-Study- Nov-2014-final.pdf  Shepherd, P. 2012. 1958 National Child Development Study user guide: Measures of ability at ages 7 to 16. Centre for Longitudinal Studies, December 2012. https://cls.ucl.ac.uk/wp- content/uploads/2017/07/NCDS-user-guide- NCDS1-3-Measures-of-ability-P-Shepherd- December-2012.pdf |

| 16 (mother) | What age did your teenage girl have  her first menstrual period? | Before 11th birthday When aged 11  Aged 12 Aged 13 Aged 14  Aged 15 or more  Not yet commenced  Commenced but don't know age | 16 (mother) | At what age did she have her first menstrual  period? | Before 11th birthday  When aged 11 Aged 12  Aged 13 Aged 14  Aged 15 or more  Not yet commenced  Commenced but don't know age  Don't know whether commenced | Age at menarche | (in years) |  |
| --- | --- | --- | --- | --- | --- | --- | --- | --- |
| 16 (medical examinat | Height in cm, to nearest 0.5 cm Height in feet and inches, to nearest 1/4 inch  Weight in kilograms, to nearest 0.1 kg  Weight in pounds and ounces | … cms  … feet … inches  … kg  … pound … ounces | 16 (medical examina | Height in cm  Height in feet and inches  Weight | … cm  … ft … in  … kg  … st … lb. | BMI at 16 | weight [kg]/height [m]2 |  |
| 16 | Which of the following most nearly  describes you? | I have never smoked a cigarette  I have only ever tried smoking once or twice  I used to smoke sometimes, but I don't now  I smoke and I would like to give it up  I do not want to give up smoking | 16 | How many cigarettes do you usually smoke  in a week? | Less than 1 a week Between 1 and 9 a week Between 10 and 19 a week Between 20 and 29 a week Between 30 and 39 a week | Has smoked at 16 | No  Yes |  |
| 16 | If you have had any alcoholic drink since this time last week, on how many days did you do so? | {number of} days | 16 | How long is it since you had an alcoholic  drink (beer, wine, spirits, etc.)? | Less than 1 week 2-4 weeks  5-8 weeks  9-12 weeks Over 12 weeks  Uncertain/Can't remember  Never had one | Has had an alcoholic drink the week  prior the age 16 interview | No  Yes |  |
| 16 | During the past year, which of the following {team/individual/other listed} sports did you play, in school and out of school, when they were in season, and how often? | At least once a week  At least once a month | 16 | Below is a list of things which many people do in their spare time. You will probably only do a few of these. Pelase show by ringing one of the numbers for each one whether this is something that you do often, sometimes, never or hardly ever. If it is something that you would like to do but don't have the chance, please ring 4.  Playing outdoor games and sports Swimming  Playing indoor games and sports | Often/Sometimes/Never or hardly ever/  Like to but no chance | Frequency of exercise at 16 | Monthly or less often  Weekly (or Often) |  |
| 16 (mother) | Below is a series of descriptions of behavior sometimes shown by young people. Please say whether, in respect of your teenager, the descriptions certainly applies, applies somewhat of doesn't apply.  Often worried, worries about many things  Often appears misarable, unhappy, tearful or distresssed  Tends to be ferful or afraid of new  things or new situations | Doesn't apply Applies somewhat Certainly applies | 16 (mother) | Below are a series of descriptions of behaviour often shown by young people. Please ask the informant about each one and ring the appropriate number to show the degree to which this description is true of the study child.  Often worried, worries about many things Often appears misarable, unhappy, tearful or distressed  Tend to be fearful or afraid of new things or  new situations | Doesn't apply Applies somewhat Certainly applies | Emotional/Neurotic score at 16  (Rutter) | Range: 0-6 | Psychological distress in childhood derived as a score from the ‘fearful’, ‘miserable’ and ‘worried’ items, loading on the Rutter emotional-neurotic factor, and ranging from 0 to 6, where 0 means that none of these behaviours apply and 6 - that all apply. |
| 30 | How tall are you without shoes?  Before you were pregnant, what was your weight?/ What is your current weight without clothes on? | Metres and Centimetres Feet and inches  Kilgrams  Stones and pounds | 33 | How tall are you without shoes?  Before you were pregnant, what was your weight?/ What is your current weight without clothes on? | Metres and Centimetres Feet and inches  Kilgrams  Stones and pounds | BMI at 30/33 | weight [kg]/height [m]2 |  |
| 30 | Would you say that..? | You've never smoked cigarettes  You used to smoke cigarettes but don't at all now  You now smoke cigarettes occasionally but  not every day  You smoke cigarettes every day | 33 | Do you smoke cigarettes at all nowadays?  Have you ever smoked cigarettes regularly - by regularly I mean at least one cigarette a day for 12 months or more? | Yes/No  Yes/No | Has smoked at 30/33 | Never Ex-smoker Smoker |  |
| 30 | How often do you have an alcoholic drink of any kind. Would you say you had a drink..? | On most days  2 to 3 days a week Once a week  2 to 3 times a month  Less often or only on special occasions  Never now a days  Have never had an alcoholic drink | 33 | How often do you have an alcoholic drink of  any kind? | Most days  1, 2 or 3 times a week 1, 2 of 3 times a month  Less often/only on special occasions  Never | Frequency of alcohol consumption at  30/33 | Weekly (once to most days a week) Monthly (one to three times a month) Less often or never |  |

| 30 | Do you regularly take part in any of the activities on this card - that is at least once month, for most of the year?  How often do you take part in any acivity of this type? | Yes  No  Every day or most days 4-5 days a week  2-3 days a week  Once a week  2-3 times a month  Less often | 33 | Do you regularly take part in any of the activities on this card - that is at least once month, for most of the year?  How often do you take part in any acivity of this type? | Yes  No  Can't say  Every day or most days  4-5 days a week 2-3 days a week Once a week  2-3 times a month Less often  Can't say | Frequency of excercise at 30/33 | Weekly (once to most days per week) Monthly (two to three times a month) Less often or never |  |
| --- | --- | --- | --- | --- | --- | --- | --- | --- |
| 30 | Malaise inventory, 24-items measuring  psycological distress |  | 33 | Malaise inventory, 24-items measuring  psycological distress |  | Has experienced depression symptoms  (Malaise) at 30/33 | No  Yes | The measure included a score of women’s responses to the Malaise inventory (Rutter et al 1970), ranging from 0 to 24. A score of 8 or higher is considered a cut off for experiencing symptoms consistent with depression, and used here to derive a binary measure. |
| 30 | Have you ever had or been told that you had a problem with your periods?  Have you ever been told that you had  any other gynaecological problems? | Yes  No  Yes  No | 33 | Have you ever suffered from or been told you had..?  Persistent trouble with periods  Other gynaecological problems | Yes  Only when preganant  No | Has suffered periods or other  gynaecological problems by 30/33 | No  Yes |  |
| 30 | Are you currently taking the contraceptive pill?  Have you ever taken the contraceptive  pill? | Yes  No  Yes  No | 42 | Are you currently taking the contraceptive pill?  Have you ever taken the contraceptive pill? | Yes  No  Yes  No | Has ever taken contraceptive pill by  30/42 | No  Yes |  |
| 33, 38 | Has anyone you were having a sexual relationship with ever become pregnant?/ have you been pregnant?  What was the result of this pregnancy  [for the nth baby]? | Yes  No  Live birth Still birth Miscarriage Abortion  Still pregnant | 33, 42 | Has anyone you were having a sexual relationship with ever become pregnant?/ have you been pregnant?  What was the result of this pregnancy [for  the nth baby]? | Yes  No  Live birth Still birth Miscarriage Abortion  Still pregnant | Has live born children | No  Yes |  |
| 38 | What is your (main) job?  What do you mainly do in your job? What does the firm or organisation  you work for mainly make or do (at the  place where you work)?  Do you have any managerial duties, or | Open text Open text Open text  Yes/No | 42 | What is your (main) job?  What do you mainly do in your job?  What does the firm or organisation you work for mainly make or do (at the place where you work)?  Do you have any managerial duties, or are | Open text Open text Open text  Yes/No | Social class at 38/42 | Non-manual  Manual  Not working or other | For comparability with social class at birth, the Registrar-General's Social class scheme was used for the adult social class measure. |

**Table A2. Questions and response options in 1970 BCS and 1958 NCDS cohorts and new variables created following data harmonisation**

| **1970 BCS cohort** | | | **1958 NCDS cohort** | | | **Harmonised** | | |
| --- | --- | --- | --- | --- | --- | --- | --- | --- |
| **Age** | **Question** | **Categories or scale** | **Age** | **Question** | **Categories or scale** | **Variable** | **Categories** | Note |
| 42 | Have you ever had an operation to remove one or both of your ovaries? | Removal of one ovary (oophorectomy)  Removal of both ovaries (bilateral oophorectomy)  None of these | 44/45 | Have you ever had any of the following operations? IF YES, give dates of all operations. If you cannot remember the month and year, give your age at the time of the operation.  Removal of uterus (womb) and both ovaries (hysterectomy and bilateral oopherectomy)  Removal of uterus (womb) only (hysterectomy)  Removal or uterus (womb) and one ovary (hysterectomy and oopherectomy)  Removal of both ovaries only (bilateral oopherectomy) | Yes/No  Month/Year/Age | Menopause  status | Pre-menopausal Peri-menopausal Menopause <40 years Menopause 40-44 years  Menopause 45 or more years No periods (other reasons) HRT  Surgery  Never had a period Insufficient information  Did not participate in follow-up sweep | Age/Month at last period' not asked in NCDS at age 44/45 if 'no periods last 12 months'. Informarmation taken from Age  50 survey, where everyone was asked 'Age/Month  at/of last period'. |
|  | Have you ever had an operation to remove your uterus (womb)? | Yes/No |  | In the last 12 months have you had a period or menstrual bleeding? |  |  |  |  |
|  | {If 'Had an operation'} How old were you when you had the operation for  the removal of {uterus (womb)/ ovary/ovaries}? | Age |  | {If 'No periods last 12 months'} Were your periods stopped by… | … Surgery?  … Chemotherapy or radiation therapy?  … Pregnancy or breastfeeding?  … No obvious reason / menopause?  … Other reason, please specify |  |  |  |
|  | {If 'Had an operation'} In what month was this operation? | Month |  | {If 'Had periods last 12 months'} In the last 3 months have you had a period or  menstrual bleeding? | Yes/No |  |  |  |
|  | In the last 12 months have you had a period or menstrual bleeding? | Yes  No  Never had a period |  | On what date did your last period start?  (Include current period if bleeding now) | Day/Month/Year/Age |  |  |  |
|  | {If 'No periods last 12 months'} What was the main reason your periods  stopped? | No obvious reason /menopause Pregnancy or breast feeding Surgery  Chemotherapy or radiation therapy  Other |  | In the last few years (in the years before your last period) did your periods… | … become more regular?  … become less regular?  … remain about the samei.e. as regular/irregular as  before)? |  |  |  |
|  | In the last 3 months have you had a period or menstrual bleeding? | Yes  No |  | Have you ever had hormone replacement therapy (HRT)? | Yes/No |  |  |  |
|  | {If 'No periods last 12 months'} How old were you when you had your last  period[(however long ago this was)]? | Age |  | {If 'ever had' HRT} When did you first start HRT? | Month/Year/Age |  |  |  |
|  | {If 'No periods last 12 months'} And what was the month of your  last period? | Month |  | {If 'ever had' HRT} Before you started HRT had your menstrual periods stopped? | Yes/No |  |  |  |
|  | Please tell us about [any recent changes to your menstrual periods. In the last few years have your periods…/ the changes before your last period.  In the years before your last period did your periods...] | Become more regular  Become less regular  Remain about the same (i.e. as regular/irregular  as before) |  | {IF 'periods stopped before HRT start'} What was the date of your last period  before starting HRT? | Month/Year/Age |  |  |  |
|  | Are you currently on hormone replacement therapy (HRT)? | Yes  No |  | {If 'ever had' HRT} Are you currently taking HRT? | Yes/No |  |  |  |
|  | {If 'Not currently on HRT'} Have you ever had HRT? | Yes  No | 50 | Since [date of biomedical interview] have you / Have you ever] had an operation  to remove one or both of your ovaries? | Removal of one ovary (oophorectomy)  Removal of both ovaries (bilateral oophorectomy)  None of these |  |  |  |
|  | {If currently or ever on HRT'} How old were you when you first started  HRT? | Age |  | In [Year of biomedical interview], you told us that you had [an ovary removed (oophorectomy) / your uterus (womb) and one ovary removed (hysterectomy and oophorectomy)]. [Since [date of biomedical interview] have you / Have you ever] had a further operation to remove your remaining ovary (oophorectomy)? | Yes/No |  |  |  |
|  | {If currently or ever on HRT'} In what month did you first start HRT? | Month |  | [Since [date of biomedical interview] have you / Have you ever] had an  operation to remove your uterus (womb)? | Yes/No |  |  |  |
|  | {If currently or ever on HRT'} Before you started HRT had your menstrual  periods stopped? | Yes  No |  | {If 'Had an operation'} How old were you when you had the operation for the  removal of {uterus (womb)/ovary/ovaries}? | Age |  |  |  |
|  | {If 'periods stopped before HRT start'} How old were you when you had  your last period before starting HRT? | Age |  | {If 'Had an operation'} In what month was this operation? | Month |  |  |  |
|  | {If 'periods stopped before HRT start'} In what month was your last period  before starting HRT? | Month |  | In the last 12 months have you had a period or menstrual bleeding? | Yes/No |  |  |  |
| 46 | {Since {date last interview} have you/Have you ever} had an operation to  remove one or both of your ovaries? | Removal of one ovary (oophorectomy)  Removal of both ovaries (bilateral oophorectomy)  Neither of these |  | {If 'No periods last 12 months'} What was the main reason your periods  stopped? | No obvious reason /menopause Pregnancy or breast feeding Surgery  Chemotherapy or radiation therapy  Other |  |  |  |
|  | In {date last interview} you told us that you had had one of your ovaries removed. Since then have you had a further operation to remove your remaining ovary? | Yes  No |  | {If 'Had periods last 12 months} In the last 3 months have you had a period or  menstrual bleeding? | Yes/No |  |  |  |
|  | {Since {date last interview} have you / Have you ever} had an operation to  remove your uterus (womb)? | Yes  No |  | What age were you when you started your last period[(however long ago this  was)]? | Age |  |  |  |
|  | {If 'Had an operation'} How old were you when you had the operation for  the removal of {uterus (womb)/ovary/ovaries}? | Age |  | And what was the month of your last period? | Month |  |  |  |

|  | {If 'Had an operation'} In what month was this operation? | Month |  | Please tell us about the [most recent changes to your menstrual periods. In the last few years have your periods…/ changes before your last period. In the years before your last period did your periods...] | ...become more regular? ...become less regular?  ...remain about the same (i.e. as regular/irregular as  before)? |
| --- | --- | --- | --- | --- | --- |
|  | In the last 12 months have you had a period or menstrual bleeding? | Yes/No |  | Are you currently on hormone replacement therapy (HRT)? | Yes/No |
|  | {If 'No periods last 12 months'} What was the main reason your periods  stopped? | No obvious reason /menopause Pregnancy or breast feeding Surgery  Chemotherapy or radiation therapy  Other |  | {If 'Not currently on HRT'} Have you ever had HRT? | Yes/No |
|  | {If 'Had periods last 12 months} In the last 3 months have you had a  period or menstrual bleeding? | Yes/No |  | {If currently or ever on HRT'} What age were you when you first started HRT? | Age |
|  | {If 'No periods last 12 months'} How old were you when you had your  last period? | Age |  | {If currently or ever on HRT'} In what month did you first start HRT? | Month |
|  | {If 'No periods last 12 months'} And what was the month of your last  period? | Month |  | {If currently or ever on HRT'} Before you started HRT had your menstrual periods  stopped? | Yes/No |
|  | Please tell us about {any recent changes to your menstrual periods. In the last few years have your periods…/ the changes before your last period. In the years before your last period did your periods...} | Become more regular Become less regular  Remain about the same (i.e. as regular/irregular  as before) |  | {If 'periods stopped before HRT start'} What age were you when you had your  last period before starting HRT? | Age |
|  | Are you currently on hormone replacement therapy (HRT)? | Yes/No |  | {If 'periods stopped before HRT start'} In what month was your last period  before starting HRT? | Month |
|  | {If 'Not currently on HRT'} Have you ever had HRT? | Yes/No |  |  |  |
|  | {If currently or ever on HRT'} How old were you when you first started  HRT? | Age |  |  |  |
|  | {If currently or ever on HRT'} In what month did you first start HRT? | Month |  |  |  |
|  | {If currently or ever on HRT'} Before you started HRT had your menstrual  periods stopped? | Yes/No |  |  |  |
|  | {If 'periods stopped before HRT start'} How old were you when you had  your last period before starting HRT | Age |  |  |  |
|  | {If 'periods stopped before HRT start'} In what month was your last period  before starting HRT? | Month |  |  |  |

**Table A2.1 Classification of women into menopause status in 1970 BCS cohort**	**Table A2.2 Classification of women into menopause status in 1958 NCDS cohort**

**1970 BCS cohort**	**1958 NCDS cohort**

**Menopause status**	**Age 42**	**Age 46**	**Menopause status**	**Age 44/45**	**Age 50**

n	%	n	%	n	%	n	%

Menopause <40 years

(premature menopause)

44	0.9

53	1.2

Menopause <40 years

(premature menopause)

131	2.8	176	3.5

Menopause 40-44 years (early menopause)

86	1.7	169	3.7

Menopause 40-44 years (early menopause)

106	2.2	207	4.1

Menopause 45 or more years	n.a.	123	2.7	Menopause 45 or more years	23	0.5	797	15.6

Peri-menopause	812	15.9	1,090	24.0	Peri-menopause	841	17.8 1,440	28.2

Pre-menopause	3,390	66.2	1,938	42.7	Pre-menopause	2,833	60.1 1,129	22.1

HRT (before FMP)	72	1.4	176	3.9	HRT (before FMP)	268	5.7	542	10.6

Hysterectomy/Bilateral oophorectomy (before FMP)

182	3.6	331	7.3

Hysterectomy/Bilateral oophorectomy (before FMP)

294	6.2	535	10.5

No periods (other reasons)	414	8.1	241	5.3	No periods (other reasons)	108	2.3	197	3.9

Never had a period	31	0.6	31	0.7	Never had a period	-	-

Insufficient information	86	1.7 384 [1]	8.5	Insufficient information	108	2.3	75	1.5

Not in age 46 survey	878	Not in age 44/45 survey	411

Total	5,117	100.0	4,536	100.0	Total	4,712	100.0 5,098	100.0

*Notes:*

*Overall number of women (and denominator) excludes cases non- participating in subsequent sweep (i.e. 878 women who took part at age 42 but not age 46).*

*Notes:*

*Overall number of women (and denominator) excludes cases non-participating in subsequent sweep (i.e. 411 women who took part at age 44/45 but not age 50).*

*Women who have undergone hysterectomy or bilateral oophorectomy prior to their FMP or whose periods stopped for other obvious reasons (e.g. pregnancy, contraceptives, chemotherapy or radiotherapy), as well as women who started HRT prior to the FMP, were excluded from this*

*analysis.*

*Women who have undergone hysterectomy or bilateral oophorectomy prior to their FMP or whose periods stopped for other obvious reasons (e.g. pregnancy, contraceptives, chemotherapy or radiotherapy), as well as women who started HRT prior to the FMP, were excluded from this analysis.*

*Analytical sample further excludes twins and triplets and women with missing multiple birth information, retaining only singletons.*

*Analytical sample further excludes twins and triplets and women with missing multiple birth information, retaining only singletons.*

*[1] Due to an error, women at age 46 in the 1970 cohort who at age 42 reported no periods in the past 12 months for reasons different from natural menopause, including pregnancy and contraceptive use, were not asked whether they had periods in the past 12 months, resulting in insufficient information to determine their menopause status at age 46 survey.*

**Table A3. Birth, childhood, and early adulthood characteristics of pre-, peri- and post-menopausal women in 1970 BCS and 1958 NCDS cohorts**

| **Characteristic** | **1970 BCS cohort** | | | | **1958 NCDS cohort** | | | |
| --- | --- | --- | --- | --- | --- | --- | --- | --- |
|  | n | % |  | | n | % |  |  |
| **Menopausal status** | **3,191** | **100.0** |  |  | **3,614** | **100.0** |  |  |
| Pre-, peri- or postmenopausal 45 or more years | 2,985 | 93.5 |  |  | 3,246 | 89.8 |  |  |
| Menopause before 45 years (Early menopause) | 206 | 6.5 |  |  | 368 | 10.2 |  |  |
| **Birth** | n | % | 95% CI | | n | % | 95% CI |  |
| **Birthweight standardized for gestation** | **2,928** | **100.0** |  |  | **3,024** | **100.0** |  |  |
| 1st quarter (lowest) | 836 | 28.6 | 26.9 | 30.2 | 886 | 29.3 | 27.7 | 30.9 |
| 2nd quarter | 785 | 26.8 | 25.2 | 28.4 | 840 | 27.8 | 26.2 | 29.4 |
| 3rd quarter | 716 | 24.5 | 22.9 | 26.0 | 696 | 23.0 | 21.5 | 24.6 |
| 4th quarter (highest) | 591 | 20.2 | 18.8 | 21.7 | 602 | 19.9 | 18.5 | 21.4 |
| *missing* | *263* | *8.2* |  |  | *590* | *16.3* |  |  |
| **Maternal age** | **3,087** | **100.0** |  |  | **3,465** | ***100.0*** |  |  |
| 19 or less years | 263 | 8.5 | 7.6 | 9.6 | 179 | 5.2 | 4.5 | 6.0 |
| 20-34 years | 2,566 | 83.1 | 81.8 | 84.4 | 2,852 | 82.3 | 81.0 | 83.5 |
| 35 or more years | 258 | 8.4 | 7.4 | 9.4 | 434 | 12.5 | 11.5 | 13.7 |
| *missing* | *104* | *3.3* |  |  | *149* | *4.1* |  |  |
| **Mother smoked during pregnancy** | **3,075** | **100.0** |  |  | **3,422** | ***100.0*** |  |  |
| No | 1,751 | 56.9 | 55.2 | 58.7 | 2,060 | 60.2 | 58.5 | 61.8 |
| Stopped prior or during the pregnancy | 153 | 5.0 | 4.3 | 5.8 | 249 | 7.3 | 6.5 | 8.2 |
| Smoked during pregnancy | 1,171 | 38.1 | 36.4 | 39.8 | 1,113 | 32.5 | 31.0 | 34.1 |
| *missing* | *116* | *3.6* |  |  | *192* | *5.3* |  |  |
| **Father's social class** | **3,075** | **100.0** |  |  | **3,467** | **100.0** |  |  |
| No father in household/Other | 206 | 6.7 | 5.9 | 7.6 | 166 | 4.8 | 4.1 | 5.6 |

| Manual | 1,873 | 60.9 | 59.2 | 62.6 | 2,297 | 66.3 | 64.7 | 67.8 |
| --- | --- | --- | --- | --- | --- | --- | --- | --- |
| Non-manual | 996 | 32.4 | 30.8 | 34.1 | 1,004 | 29.0 | 27.5 | 30.5 |
| *missing* | *116* | *3.6* |  |  | *147* | *4.1* |  |  |
| **Breastfed (5/7)** | **2,663** | **100.0** |  |  | **3,190** | **100.0** |  |  |
| Less than 1 month or never | 1,990 | 74.7 | 73.0 | 76.3 | 1,704 | 53.4 | 51.7 | 55.1 |
| 1 or more months | 673 | 25.3 | 23.7 | 27.0 | 1,486 | 46.6 | 44.9 | 48.3 |
| *missing* | *528* | *16.6* |  |  | *424* | 11.7 |  |  |
| **Childhood** |  |  |  |  |  |  |  |  |
| **Cognitive ability score at 10/11 (maths, reading)** | **2,473** |  |  |  | **3,142** |  |  |  |
| mean (s.d.) | 0.2 (0.9) |  |  |  | 0.2 (0.9) |  |  |  |
| median (range) | 0.2 (-3.1, 2.2) | |  |  | 0.2 (-2.2, 2.6) | |  |  |
| *missing* | *718* | *22.5* |  |  | *472* | *13.1* |  |  |
| **Age at menarche (mother's report at 16)** | **2,039** |  |  |  | **2,722** |  |  |  |
| mean (s.d.) | 12.7 (1.3) |  |  |  | 12.8 (1.3) |  |  |  |
| median (range) | 13 (10,16) |  |  |  | 13 (10,16) |  |  |  |
| *missing* | *1,152* | *36.1* |  |  | *892* | *25.7* |  |  |
| **BMI at 16** | **1,944** |  |  |  | **2,494** |  |  |  |
| mean (s.d.) | 21.3 (3.1) |  |  |  | 21.1 (3.0) |  |  |  |
| median (range) | 20.8 (9.9,47.7) | |  |  | (14.5,40.4 |  |  |  |
| *missing* | 1,247 | 39.1 |  |  | 1,120 | 31.0 |  |  |
| **Smoking at 16** | **1,432** | **100.0** |  |  | **2,805** | **100.0** |  |  |
| No | 1157 | 80.8 | 78.7 | 82.8 | 1,896 | 67.6 | 65.8 | 69.3 |
| Yes | 275 | 19.2 | 17.2 | 21.3 | 909 | 32.4 | 30.7 | 34.2 |
| *missing* | 1,759 | 55.1 |  |  | 809 | 22.4 |  |  |

| **Had a drink of alcohol**  **in week prior Age 16 interview** | **1,379** | **100.0** |  |  | **2,808** | **100.0** |  |  |
| --- | --- | --- | --- | --- | --- | --- | --- | --- |
| No | 457 | 33.1 | 30.7 | 35.7 | 1,654 | 58.9 | 57.1 | 60.7 |
| Yes | 922 | 66.9 | 64.3 | 69.3 | 1,154 | 41.1 | 39.3 | 42.9 |
| *missing* | *1,812* | *56.8* |  |  | *806* | 22.3 |  |  |
| **Exercise at 16** | **1,450** | 100.0 |  |  | **2,757** | **100.0** |  |  |
| Less often | 169 | 11.7 | 10.1 | 13.4 | 1,678 | 60.9 | 59.0 | 62.7 |
| Weekly/Often | 1,281 | 88.3 | 86.6 | 89.9 | 1,079 | 39.1 | 37.3 | 41.0 |
| *missing* | *1,741* | *54.6* |  |  | *857* | *23.7* |  |  |
| **Emotional/Neurotic score at 16 (Rutter)** | **2,087** |  |  |  | **2,749** |  |  |  |
| mean (s.d.) | 1.1 (1.2) |  |  |  | 1.1 (1.2) |  |  |  |
| median (range) | 1 (0,6) |  |  | | 1 (0,6) |  |  |  |
| *missing* | *1,104* | 34.6 |  |  | *865* | 23.9 |  |  |
| **Adulthood** |  |  |  |  |  |  |  |  |
| **BMI at 30/33** | **2,652** |  |  |  | **2,874** |  |  |  |
| mean (s.d.) | 24.0 (4.5) |  |  |  | 24.4 (4.8) |  |  |  |
| median (range) | 23.05 (14.9,58.2) | |  |  | 23.3 (11.3,50.2) | |  |  |
| *missing* | *539* | *16.9* |  |  | *740* | *20.5* |  |  |
| **Smoking at 30/33** | **2,836** | **100.0** |  |  | **3,208** | **100.0** |  |  |
| Never | 1,328 | 46.8 | 45.0 | 48.7 | 1,704 | 53.1 | 51.4 | 54.8 |
| Ex-smoker | 590 | 20.8 | 19.3 | 22.3 | 593 | 18.5 | 17.2 | 19.9 |
| Smoker | 918 | 32.4 | 30.7 | 34.1 | 911 | 28.4 | 26.9 | 30.0 |
| *missing* | *355* | *11.1* |  |  | 406 | 11.2 |  |  |
| **Frequency of alcohol consumption at 30/33** | **2,836** | **100.0** |  |  | **3,220** | **100.0** |  |  |
| Less often or never | 662 | 23.3 | 21.8 | 24.9 | 883 | 27.4 | 25.9 | 29.0 |
| Monthly (one to three times a month) | 450 | 15.9 | 14.6 | 17.3 | 771 | 23.9 | 22.5 | 25.4 |
| Weekly (once to most days a week) | 1,724 | 60.8 | 59.0 | 62.6 | 1,566 | 48.6 | 46.9 | 50.4 |

| *missing* | *355* | *11.1* |  |  | *394* | *10.9* |  |  |
| --- | --- | --- | --- | --- | --- | --- | --- | --- |
| **Exercise at 30/33** | **2,835** | **100.0** |  |  | **3,209** | **100.0** |  |  |
| Less often or never | 675 | 23.8 | 22.28 | 25.41 | 767 | 23.9 | 22.5 | 25.4 |
| Monthly (two to three times a month) | 134 | 4.7 | 4.00 | 5.57 | 183 | 5.7 | 5.0 | 6.6 |
| Weekly (once to most days per week) | 2,026 | 71.5 | 69.77 | 73.10 | 2,259 | 70.4 | 68.8 | 72.0 |
| *missing* | *356* | *11.2* |  |  | *405* | *11.2* |  |  |
| **Experienced depression symptoms (Malaise) at 30/33** | **2,821** | **100.0** |  |  | **3,214** | **100.0** |  |  |
| No | 2,460 | 87.2 | 85.9 | 88.4 | 2,974 | 92.5 | 91.6 | 93.4 |
| Yes | 361 | 12.8 | 11.6 | 14.1 | 240 | 7.5 | 6.6 | 8.4 |
| *missing* | *370* | *11.6* |  |  | *400* | 11.1 |  |  |
| **Suffered period or other gynaecological problems** (by age 30/33) | **2,837** | **100.0** |  |  | **3,190** | **100.0** |  |  |
| No | 2,037 | 71.8 | 70.1 | 73.4 | 2,400 | 75.2 | 73.7 | 76.7 |
| Yes | 800 | 28.2 | 26.6 | 29.9 | 790 | 24.8 | 23.3 | 26.3 |
| *missing* | *354* | *11.1* |  |  | *424* | *11.7* |  |  |
| **Use of contraceptives** (by age 30/42) | **2,837** | **100.0** |  |  | **3,431** | **100.0** |  |  |
| No | 257 | 9.1 | 8.1 | 10.2 | 352 | 10.3 | 9.3 | 11.3 |
| Yes | 2,580 | 90.9 | 89.8 | 91.9 | 3,079 | 89.7 | 88.7 | 90.7 |
| *missing* | *354* | *11.1* |  |  | *183* | *5.1* |  |  |
| **Nulliparous (no live births)** (by age 38/42) | **2,768** | **100.0** |  |  | **3,244** | **100.0** |  |  |
| No | 1,985 | 71.7 | 70.0 | 73.4 | 2,718 | 83.8 | 82.48 | 85.01 |
| Yes | 783 | 28.3 | 26.6 | 30.0 | 526 | 16.2 | 14.99 | 17.52 |
| *missing* | *423* | *13.3* |  |  | *370* | *10.2* |  |  |
| **Social class 3 classes** (age 38/42) | **2,675** | **100.0** |  |  | **3,430** | **100.0** |  |  |
| Not working/Other | 570 | 21.3 | 19.8 | 22.9 | 680 | 19.8 | 18.52 | 21.19 |
| Manual | 403 | 15.1 | 13.8 | 16.5 | 725 | 21.1 | 19.80 | 22.54 |
| Non-manual | 1,702 | 63.6 | 61.8 | 65.4 | 2,025 | 59.0 | 57.38 | 60.67 |

| *missing* | *516* | *16.2* |  |  | *184* | *5.1* |
| --- | --- | --- | --- | --- | --- | --- |

Note: Missing values excluded from analysis

**Table A4. Odds Ratios (OR) and 95% confidence intervals (CI) for being early menopausal on birth, childhood and adult characteristics (imputed, n=6,805)**

**RISK FACTORS**	**BIRTH**	**CHILDHOOD**	**ADULTHOOD**

Crude/Unadjusted	Adjusted	Adjusted	Adjusted

**OR**

**95% CI**

**OR**

**95% CI**

**OR**

**95% CI**

**OR**

**95% CI**

**Maternal age /**

**Birthweight**

**OR**

**95% CI**

**OR**

**95% CI**

**Age at menarche /**

**BMI / Smoking / Drinking /**

**OR**	**95% CI**

**BMI / Smoking /**

**Drinking / Exercise / Emotional**

**distress / Gynaecological**

**Father's social class**

**Mother smoked during pregnancy**

**standardized for**

**gestation**

**Breastfed**

**Cognitive ability score (maths & reading) at 10/11**

**Exersice / Emotional distress at**

**16**

**problems / Contraceptives /**

**Live birth / Social class**

**BIRTH**

**Birthweight standardized for gestation**

1st quarter (lowest)	1.09	0.85	1.40

1.01	0.78	1.30

1.00	0.77	1.28

0.92	0.71	1.19

0.93	0.72	1.20

0.93	0.71	1.20

2nd quarter	0.94	0.73	1.22	0.91	0.70	1.18	0.91	0.70	1.18	0.90	0.69	1.17	0.90	0.69	1.18	0.92	0.70	1.21

3rd quarter (reference)

4th quarter (highest)	1.03	0.79	1.36

1.05	0.80	1.39

1.05	0.80	1.38

1.05	0.80	1.39

1.02	0.77	1.36

1.05	0.79	1.39

**Maternal age at birth**

19 or less years 1.39*	1.00	1.92	1.25	0.90	1.74	1.24	0.89	1.73	1.14	0.81	1.59	1.12	0.80	1.56	1.11	0.79	1.55

20-34 years (reference)

35 or more years	1.10	0.84	1.45

1.13	0.85	1.48

1.11	0.85	1.47

1.15	0.87	1.52

1.16	0.87	1.53

1.15	0.87	1.54

**Mother smoked during pregnancy**

No (reference)

Stopped prior or during the pregnancy	0.79	0.52	1.20

0.77	0.50	1.18

0.76	0.50	1.17

0.76	0.50	1.17

0.78	0.51	1.19

0.74	0.48	1.14

0.70	0.45	1.08

Smoked during pregnancy 1.32**	1.10	1.59	**1.24***	**1.03**	**1.49**	1.24*	1.03	1.49	1.21*	1.00	1.46	1.12	0.93	1.36	1.07	0.88	1.30	1.02	0.84	1.25

**Father's social class**

No father in household/Other 2.20***	1.48	3.28	**2.20*****	**1.48**	**3.28**	2.12***	1.42	3.17	2.05***	1.36	3.08	1.95***	1.29	2.94	1.55*

1.02	2.35	1.48*

0.98	2.26	1.50*

0.98	2.29

Manual 2.02***	1.61	2.54	**2.02*****	**1.61**	**2.54**	1.97***	1.57	2.48	1.96***	1.56	2.46	1.90***	1.51	2.39	1.48**	1.17	1.88	1.45***	1.14	1.84	1.44***	1.13	1.84

Non-manual (reference)

**Breastfed**

Less than 1 month or never 1.44***	1.18	1.77	**1.30***	**1.05**	**1.60**	1.20+	0.97	1.48	1.21*	0.98	1.50	1.20	0.97	1.49

Breastfed 1 or more months (reference)

**CHILDHOOD**

**Cognitive ability score (maths & reading) at**

0.59	0.54	0.66

**0.64*****

**0.57**	**0.71**	0.65***

0.58	0.73	0.69***

0.62	0.78

**Age at menarche (per year)**	0.97	0.90	1.05	0.98	0.90	1.07	0.97	0.89	1.06

**BMI at 16 (per kg/m2)**	1.05***	1.01	1.09	1.03	0.99	1.07	1.03	0.98	1.09

**Smoking at 16**

No (reference)

Yes 1.76***	1.41	2.19

**1.51*****

**1.19**	**1.92**

1.12	0.82	1.54

**Drinking the week prior age 16 interview**

No (reference)

Yes	1.07	0.86	1.34

1.09	0.85	1.39

1.10	0.86	1.40

**Exercise at 16**

Monthly or less often (reference)

Weekly (or often))	0.90	0.68	1.17

0.92	0.70	1.20

0.95	0.72	1.25

**Emotional/neurotic subscore at 16 (Rutter)**	1.03	0.94	1.12	1.00	0.92	1.10	0.97	0.88	1.06

**ADULTHOOD**

**BMI at 30/33 (per kg/m2)**

1.02+	1.00	1.04

0.99	0.96	1.02

**Smoking at 30/33**

Never (reference)

Ex-smoker	1.16	0.88	1.52

1.03	0.77	1.39

Smoker 2.36***	1.94	2.88	**1.69*****	**1.28**	**2.23**

**Alcohol consumption at 30/33**

Less often or never (reference)

Monthly (one to three times a month) 0.67***	0.51	0.87

**0.76****

**0.57**	**1.00**

Weekly (once to most days a week) 0.68***	0.55	0.84	0.84	0.67	1.06

**Exercise at 30/33**

Less often or never (reference)

Monthly (two to three times a month)	1.00	0.68	1.47

1.18	0.79	1.76

Weekly (once to most days per week) 0.63***	0.51	0.78	**0.75*****	**0.60**	**0.93**

**Depressive symptoms at 30/33 (Malaise)**

No (reference)

Yes 1.76***	1.35	2.30

1.11	0.83	1.48

**Gynaecological problems by 30/33**

No (reference)

Yes 1.81***	1.48	2.21

**1.68*****

**1.36**	**2.06**

**Use oral contraceptives by 30/42**

No (reference)

Yes 0.77+	0.58	1.02

0.85	0.63	1.16

**Live born children by 38/42**

No (reference)

Yes	0.94	0.74	1.19

1.13	0.88	1.45

**Social class 3 classes at age 38/42**

Not working/Other 1.82***	1.46	2.25

**1.43*****

**1.13**	**1.81**

Manual 1.32*	1.04	1.68	0.96	0.74	1.24

Non-manual (reference)

**Cohort year**

1958

1970 (reference)

All univariate models account for cohort year

1.62***	1.35	1.93	1.65***	1.38	1.98	1.65***	1.38	1.98	1.75***	1.45	2.10	1.72***

1.42	2.07	1.62***

1.27	2.06	1.81***

1.41	2.34

Note: An asterisk indicates significance levels, i.e. *** p<0.001, ** p<0.01, * p<0.05, +p<0.1

**Table A5. Odds Ratios (OR) and 95% confidence intervals (CI) for being early menopausal on birth, childhood and adult characteristics in 1970 BCS cohort (n=3,191)**

**Complete case**	**Imputed (MI)**

**RISK FACTORS**	**BIRTH**	**CHILDHOOD**	**ADULTHOOD**

Crude/Unadjusted	Crude/Unadjusted	Adjusted	Adjusted	Adjusted

OR

95% CI

OR

95% CI

OR

95% CI

OR

95% CI

OR

95% CI

**Maternal age /**

OR

95% CI

OR

95% CI

OR

95% CI

**Age at menarche /**

OR

95% CI

**BMI / Smoking /**

**Father's social class**

**Mother smoked during pregnancy**

**Birthweight standardized for**

**gestation**

**Breastfed**

**Cognitive ability score (maths & reading) at 10/11**

**BMI / Smoking / Drinking / Exersice / Emotional distress at**

**Drinking / Exercise / Emotional distress / Gynaecological problems**

**BIRTH**

**Birthweight standardized for gestation**

1st quarter (lowest)	1.12	0.74	1.68

1.12	0.75	1.67

1.05	0.70	1.57

1.00	0.67	1.50

0.90	0.60	1.36

0.88	0.57	1.34

0.90	0.58	1.38

2nd quarter	1.04	0.68	1.58	1.03	0.68	1.57	1.00	0.66	1.53	1.00	0.66	1.53	0.99	0.64	1.51	0.98	0.63	1.53	1.01	0.65	1.58

3rd quarter (reference)

4th quarter (highest)	1.05	0.67	1.64

1.04	0.66	1.62

1.07	0.68	1.67

1.04	0.67	1.64

1.03	0.65	1.63

1.01	0.63	1.60

1.04	0.65	1.67

**Maternal age at birth**

19 or less years 1.62*	1.04	2.53	1.59*	1.02	2.48	1.45	0.92	2.30	1.41	0.89	2.23	1.25	0.79	1.99	1.22	0.75	1.98	1.18	0.72	1.94

20-34 years (reference)

35 or more years	1.09	0.65	1.83

1.08	0.65	1.82

1.12	0.67	1.88

1.12	0.66	1.89

1.08	0.63	1.83

1.14	0.66	1.95

1.15	0.66	1.98

**Mother smoked during pregnancy**

No (reference)

Stopped prior or during the pregnancy	0.92	0.44	1.93

0.93	0.44	1.94

0.91	0.43	1.90

0.86	0.41	1.81

0.87	0.41	1.83

0.88	0.41	1.88

0.80	0.37	1.74

0.74	0.34	1.62

Smoked during pregnancy 1.39*	1.03	1.87	1.37*	1.02	1.84	**1.30+**	**0.96**	**1.75**	1.29	0.95	1.74	1.20	0.88	1.63	1.06	0.78	1.45	0.97	0.70	1.34	0.93	0.67	1.30

**Father's social class**

No father in household/Other 1.86*

1.04	3.31	**1.84***	**1.03**	**3.27**	**1.84***	**1.03**	**3.27**	1.75+

0.98	3.12

1.60	0.88	2.91

1.43	0.79	2.61

1.08	0.58	2.00

1.02	0.54	1.90

1.05	0.56	1.98

Manual 1.59**	1.13	2.25	**1.57***	**1.11**	**2.21**	**1.57***	**1.11**	**2.21**	1.52*	1.07	2.14	1.49*	1.05	2.10	1.34	0.94	1.90	0.97	0.67	1.40	0.92	0.63	1.33	0.91	0.62	1.33

Non-manual (reference)

**Breastfed (5)**

Less than 1 month or never 2.66***	1.64	4.34	2.66***	1.65	4.28	**2.40*****	**1.48**	**3.90**	**2.12*****	**1.30**	**3.46**	2.16**	1.32	3.55	2.21**	1.27	3.49

Breastfed 1 or more months (reference) **CHILDHOOD**

**Cognititve ability score at 10 (maths, readin**

0.52***	0.43	0.61	0.51***	0.43	0.60

**0.53*****	**0.44**	**0.63**	0.55***	0.46	0.66	0.57***

0.47	0.69

**Age at menarche (mother's report at 16) (pe**	0.90	0.78	1.04	0.90	0.78	1.04	0.93	0.79	1.09	0.92	0.78	1.08

**BMI at 16 (per kg/m2)**	1.08**	1.03	1.14	1.09**	1.03	1.15	**1.06***	**1.00**	**1.13**	1.08+	1.00	1.17

**Smoking at 16**

No (reference)

Yes 1.73*

1.05	2.86	2.10**	1.32	3.34

**1.84***

**1.10**	**3.09**

1.46	0.73	2.91

**Drinking the week prior age 16 interview**

No (reference)

Yes	0.88	0.55	1.42

1.05	0.69	1.59

0.95	0.59	1.53

0.93	0.57	1.54

**Exercise at 16**

Monthly or less often (reference)

Weekly (or often)	0.69	0.38	1.28

0.64	0.33	1.24

0.73	0.37	1.45

0.76	0.37	1.55

**Emotional/neurotic score (Rutter) at 16**

1.14+

1.00	1.30	1.14+	0.99	1.30

1.09	0.94	1.26

1.08	0.93	1.25

**ADULTHOOD**

**BMI at 30 (per kg/m2)**

1.03*	1.00	1.07	1.03+	1.00	1.06

0.98	0.94	1.03

**Smoking at 30**

Never (reference)

Ex-smoker	1.11	0.71	1.74

1.14	0.73	1.78

1.08	0.66	1.78

Smoker	2.23***	1.59	3.12	2.2***	1.57	3.10	1.49	0.89	2.49

**Alcohol consumption at 30**

Less often or never (reference)

Monthly (one to three times a month) 0.57*

0.35	0.94	0.60*	0.37	0.98

0.72	0.43	1.23

Weekly (once to most days a week) 0.65*	0.47	0.92	0.65*	0.47	0.92	1.01	0.69	1.48

**Exercise at 30**

Less often or never (reference)

Monthly (two to three times a month)	0.99	0.52	1.89

0.97	0.51	1.83

1.24	0.62	2.46

Weekly (once to most days per week) 0.56**	0.40	0.78	0.56***	0.40	0.77	**0.64***	**0.45**	**0.91**

**Depressive symptoms (Malaise) at 30**

No (reference)

Yes	1.63*	1.10	2.42	1.61*	1.09	2.39

0.99	0.63	1.54

**Gynaecological problems** (by age 30)

No (reference)

Yes	1.23	0.89	1.70

1.22	0.88	1.68

1.25	0.88	1.77

**Use of contraceptives** (by age 30)

No (reference)

Yes	0.78	0.48	1.27

0.76	0.47	1.25

0.82	0.47	1.43

**Nulliparous (no live births)** (by age 38)

No (reference)

Yes	0.89	0.63	1.26

0.90	0.64	1.28

1.17	0.78	1.75

**Social class 3 classes** (age 38)

Not working/Other	1.78**	1.22	2.61	1.90**	1.32	2.76

**1.46+**

**0.96**	**2.20**

Manual	1.98**	1.31	3.00	1.86**	1.23	2.82	1.37	0.87	2.14

Non-manual (reference)

Note: An asterisk indicates significance levels, i.e. *** p<0.001, ** p<0.01, * p<0.05, p<0.1

**Table A6. Odds Ratios (OR) and 95% confidence intervals (CI) for being early menopausal on birth, childhood and adult characteristics in 1958 NCDS cohort (n=3,416)**

**Complete case**	**Imputed (MI)**

**RISK FACTORS**	**BIRTH**	**CHILDHOOD**	**ADULTHOOD**

**Characterisitc**	Crude/Unadjusted	Crude/Unadjusted	Adjusted	Adjusted	Adjusted

OR	(95% CI)

OR

(95% CI)

OR

95% CI

OR

95% CI

OR

95% CI

**Maternal age /**

OR

95% CI

OR

95% CI

OR

95% CI

**Age at menarche /**

OR

95% CI

**BMI / Smoking /**

**Father's social class**

**Mother smoked during pregnancy**

**Birthweight standardized for**

**gestation**

**Breastfed**

**Cognitive ability score (maths & reading) at 10/11**

**BMI / Smoking / Drinking / Exersice / Emotional distress at**

**Drinking / Exercise / Emotional**

**distress / Gynaecological**

**BIRTH**

**Birthweight standardized for gestation**

1st quarter (lowest)	1.07	0.77	1.48

1.07	0.78	1.48

0.98	0.71	1.36

0.98	0.71	1.36

0.93	0.67	1.28

0.94	0.67	1.30

0.92	0.66	1.29

2nd quarter	0.86	0.61	1.21	0.89	0.64	1.24	0.86	0.62	1.20	0.86	0.62	1.20	0.85	0.61	1.19	0.85	0.61	1.20	0.89	0.63	1.26

3rd quarter (reference)

4th quarter (highest)	1.05	0.73	1.50

1.03	0.73	1.46

1.05	0.74	1.49

1.05	0.74	1.48

1.05	0.74	1.49

1.03	0.72	1.47

1.06	0.74	1.53

**Maternal age at birth**

19 or less years	1.23	0.76	1.96	1.20	0.75	1.94	1.07	0.66	1.75	1.07	0.66	1.74	1.01	0.62	1.64	0.99	0.61	1.61	1.02	0.62	1.68

20-34 years (reference)

35 or more years	1.12	0.81	1.55

1.11	0.80	1.53

1.13	0.81	1.56

1.12	0.81	1.56

1.16	0.84	1.62

1.16	0.83	1.62

1.16	0.83	1.63

**Mother smoked during pregnancy**

No (reference)

Stopped prior or during the pregnancy	0.71	0.42	1.19

0.73	0.44	1.22

0.72	0.43	1.19

0.71	0.43	1.19

0.71	0.43	1.19

0.73	0.43	1.21

0.70	0.42	1.18

0.67	0.40	1.13

Smoked during pregnancy 1.30*	1.03	1.65	1.30*	1.03	1.64	1.20	0.95	1.52	1.21	0.96	1.54	1.20	0.95	1.53	1.14	0.90	1.45	1.11	0.87	1.41	1.06	0.82	1.36

**Father's social class**

No father in household/Other 2.46**	1.43	4.23

**2.52****	**1.46**	**4.36**	**2.52****	**1.46**	**4.36**	2.43***	1.40	4.21	2.42***

1.39	4.24	2.39**	1.36	4.18	1.97*	1.12	3.48	1.93*	1.09	3.42

1.94*

1.08	3.47

Manual 2.43***	1.79	3.29	**2.41*****	**1.77**	**3.27**	**2.41*****	**1.77**	**3.27**	2.36***	1.74	3.21	2.36***	1.74	3.21	2.35***	1.72	3.19	1.92***	1.40	2.65	1.91***	1.39	2.64	1.90***	1.37	2.64

Non-manual (reference)

**Breastfed (7)**

Less than 1 month or never	1.20	0.95	1.52	1.19	0.94	1.50	1.07	0.85	1.36	1.01	0.80	1.29	1.03	0.81	1.31	1.04	0.81	1.33

Breastfed 1 or more months (reference) **CHILDHOOD**

**Cognititve ability scoreat 11 (maths, readi** 0.66***	0.58	0.76

0.66***	0.58	0.74

**0.71*****	**0.62**	**0.82**	0.72***	0.63	0.83

0.78**

0.67	0.91

**Age at menarche (mother's report at 16)v**	1.01	0.92	1.11	1.01	0.92	1.11	1.01	0.91	1.12	1.01	0.90	1.12

**BMI at 16 (per kg/m2)**	1.02	0.98	1.07	1.02	0.98	1.07	1.01	0.96	1.06	1.00	0.93	1.07

**Smoking at 16**

No (reference)

Yes 1.65***	1.28	2.12

1.60***	1.25	2.04

**1.37***	**1.06**	**1.78**

0.95	0.68	1.33

**Drinking the week prior age 16 interview**

No (reference)

Yes	1.10	0.86	1.41

1.09	0.85	1.40

1.15	0.88	1.50

1.19	0.90	1.56

**Exercise at 16**

Monthly or less often (reference)

Weekly (or often)	0.99	0.77	1.28

1.00	0.77	1.29

1.00	0.77	1.29

1.04	0.79	1.36

**Emotional/neurotic score (Rutter) at 16**

0.97	0.87	1.07

0.96	0.86	1.07

0.94	0.85	1.06

0.90+

0.80	1.01

**ADULTHOOD**

**BMI at 33 (per kg/m2)**

1.01	0.98	1.03

1.01	0.99	1.04

1.00	0.96	1.03

**Smoking at 33**

Never (reference)

Ex-smoker	1.07	0.75	1.53

1.16	0.82	1.65

1.07	0.72	1.57

Smoker 2.42***	1.87	3.12	2.46***	1.91	3.16	**1.93*****	**1.38**	**2.70**

**Alcohol consumption at 33**

Less often or never (reference)

Monthly (one to three times a month) 0.69*	0.50	0.95

0.70*	0.51	0.96

**0.73+**

**0.52**	**1.01**

Weekly (once to most days a week) 0.70**	0.54	0.92	0.70**	0.53	0.91	**0.74+**	**0.55**	**1.00**

**Exercise at 30**

Less often or never (reference)

Monthly (two to three times a month)	1.07	0.65	1.74

1.03	0.63	1.66

1.18	0.71	1.95

Weekly (once to most days per week) 0.70**	0.54	0.91	0.68**	0.53	0.89	0.82	0.62	1.08

**Depressive symptoms (Malaise) at 30**

No (reference)

Yes 1.90**	1.31	2.75

1.90**	1.32	2.74

1.22	0.82	1.82

**Gynaecological problems** (by age 33)

No (reference)

Yes 2.29***	1.80	2.92

2.27***	1.76	2.94

**2.07*****	**1.59**	**2.71**

**Use of contraceptives** (by age 42)

No (reference)

Yes	0.78	0.56	1.10

0.77	0.55	1.09

0.87	0.60	1.27

**Nulliparous (no live births)** (by age 42)

No (reference)

Yes	0.97	0.70	1.33

0.97	0.70	1.34

1.13	0.79	1.61

**Social class 3 classes** (age 42)

Not working/Other 1.78***	1.37	2.31

1.77***	1.36	2.30

**1.41***

**1.05**	**1.88**

Manual	1.10	0.82	1.47	1.11	0.83	1.48	0.81	0.59	1.11

Non-manual (reference)

Note: An asterisk indicates significance levels, i.e. *** p<0.001, ** p<0.01, * p<0.05, p<0.1
